# Characteristics and outcomes of an international cohort of 400,000 hospitalised patients with Covid-19

**DOI:** 10.1101/2021.09.11.21263419

**Authors:** ISARIC Clinical Characterisation Group, Christiana Kartsonaki

**Affiliations:** University of Oxford, UK

## Abstract

**Background:** Policymakers need robust data to respond to the COVID-19 pandemic. We describe demographic features, treatments and clinical outcomes in the International Severe Acute Respiratory and emerging Infection Consortium (ISARIC) COVID-19 cohort, the world’s largest international, standardised cohort of hospitalised patients.

**Methods:** The dataset analysed includes COVID-19 patients hospitalised between January 2020 and May 2021. We investigated how symptoms on admission, comorbidities, risk factors, and treatments varied by age, sex, and other characteristics. We used Cox proportional hazards models to investigate associations between demographics, symptoms, comorbidities, and other factors with risk of death, admission to intensive care unit (ICU), and invasive mechanical ventilation (IMV).

**Findings:** 439,922 patients with laboratory-confirmed (91.7%) or clinically-diagnosed (8.3%) SARS-CoV-2 infection from 49 countries were enrolled. Age (adjusted hazard ratio [HR] per 10 years 1.49 [95% CI 1.49-1.50]) and male sex (1.26 [1.24-1.28]) were associated with a higher risk of death. Rates of admission to ICU and use of IMV increased with age up to age 60, then dropped. Symptoms, comorbidities, and treatments varied by age and had varied associations with clinical outcomes. Tuberculosis was associated with an 86% higher risk of death, and HIV with an 87% higher risk of death. Case fatality ratio varied by country partly due to differences in the clinical characteristics of recruited patients.

**Interpretation:** The size of our international database and the standardized data collection method makes this study a reliable and comprehensive international description of COVID-19 clinical features. This is a viable model to be applied to future epidemics.

**Funding:** UK Foreign, Commonwealth and Development Office, the Bill & Melinda Gates Foundation and Wellcome. See acknowledgements section for funders of sites that contributed data.

**Research in context:** *Evidence before this study:* To identify large, international analyses of hospitalised COVID-19 patients that used standardised data collection, we conducted a systematic review of the literature from 1 Jan 2020 to 28 Apr 2020. We identified 78 studies, with data from 77,443 people (1) predominantly from China. We could not find any studies including data from low and middle-income countries. We repeated our search on 18 Aug 2021 but could not identify any further studies that met our inclusion criteria.

*Added value of this study:* Our study uses standardised clinical data collection to collect data from a vast number of patients across the world, including patients from low-, middle-, and high-income countries. The size of our database gives us great confidence in the accuracy of our descriptions of the global impact of COVID-19. We can confirm findings reported by smaller, country-specific studies and compare clinical data between countries. We have demonstrated that it is possible to collect large volumes of standardised clinical data during a pandemic of a novel acute respiratory infection. The results provide a valuable resource for present policymakers and future global health researchers.

*Implications of all the available evidence:* Presenting symptoms of SARS-CoV-2 infection in patients requiring hospitalisation are now well-described globally, with the most common being fever, cough, and shortness of breath. Other symptoms also commonly occur, including altered consciousness in older adults and gastrointestinal symptoms in younger patients, and age can influence the likelihood of a patient having symptoms that match one or more case definitions. There are geographic and temporal variations in the case fatality rate (CFR), but overall, CFR was 20.6% in this large international cohort of hospitalised patients with a median age of 60 years (IQR: 45 to 74 years).

## Introduction

To respond to COVID-19, policymakers and clinicians need robust data to drive the decision-making processes which save or cost lives. Observational cohort data describing clinical characteristics and the likelihood of severe outcomes can guide health policy development, produce research hypotheses for clinical trials and improve clinical guidelines for patient care (2).

Across the world, multiple cohort studies have described the clinical impact of the COVID-19 pandemic. However, there are difficulties with comparing diverse, fragmented datasets, which often differ in their inclusion criteria, case definitions, and outcome measures (3–9). Such heterogeneity in study design makes international comparisons challenging.

The Clinical Characterisation Protocol (CCP) developed by the International Severe Acute Respiratory and emerging Infection Consortium (ISARIC) and the World Health Organization (WHO) (10) has helped researchers across the world to collect and analyse clinical data (11). Thanks to this international cooperation, it has been possible to produce a truly global cohort study using standardised clinical data.

We present data from an international cohort of almost half a million patients from 720 sites across 49 countries. We summarise the demographic features and clinical presentation of hospitalised patients with COVID-19 in low, middle and high resource settings. We characterise the variability in clinical features in these patients and explore the risk factors associated with mortality, need for intensive care and mechanical ventilation, on a global scale.

## Methods

### Study

We used international prospective observational data of clinical features of patients admitted to hospital with COVID-19. The ISARIC/WHO CCP, incorporating Short PeRiod IncideNce sTudy of Severe Acute Respiratory Infection (SPRINT SARI) (12), is a standardised protocol for investigations of (re-)emerging pathogens of public health interest. All participating sites obtained the required local research ethics approvals; many also received a waiver of consent from the local ethics committee. All data were stored at a central repository at the University of Oxford, England. Patients with clinically suspected or laboratory-confirmed COVID-19 infection were enrolled in the study. Participating sites used the ISARIC case report form (13) to enter data onto a Research Electronic Data Capture (REDCap, version 8.11.11, Vanderbilt University, Nashville, Tenn.) database or used local databases (**Supplementary methods**) before uploading to the central data repository. Centrally collated data were converted to Study Data Tabulation Model (version 1.7, Clinical Data Interchange Standards Consortium, Austin, Tex.). The first patient was enrolled on 30 January 2020. This analysis includes all patients whose data were entered up to 24 May 2021.

### Participants

We included hospitalised patients of any age with clinical or laboratory diagnosed COVID-19. This analysis included patients admitted to hospitals in all countries contributing data. It also includes a subset of asymptomatic patients who were admitted to the hospital purely for isolation.

### Statistical analysis

We excluded patients with missing age or sex from all analyses (**Figure 1**). We additionally excluded from analyses on symptoms and treatments patients from sites that reported to have stopped collecting consistently information on symptoms and treatments. We calculated median (IQR) for continuous variables and proportions for categorical variables. We calculated proportions of patients who met each of the WHO, Centers for Disease Control and Prevention (CDC) of the United States, European Centre for Disease Prevention and Control (ECDC), and Public Health England (PHE) symptom-based case definitions (**Supplementary methods**). We calculated case fatality ratios (CFRs) overall, by country and by month, using the method suggested by Ghani et al. (14). We calculated weighted CFRs by country.

**Figure 1:**
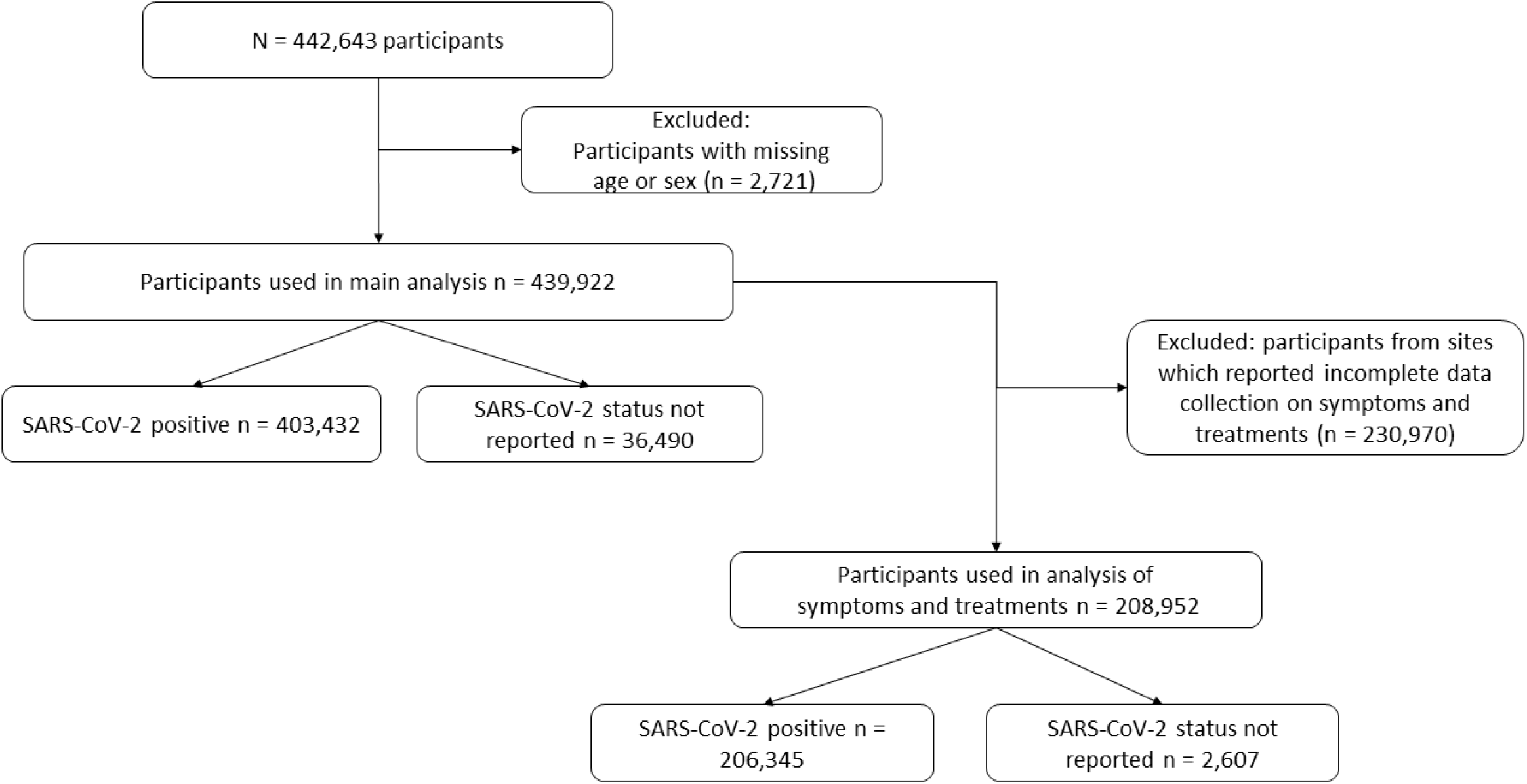
Numbers of participants.

Patients were followed up from hospital admission to death, discharge or loss to follow-up, whichever occurred first. Cox proportional hazards models were used to assess the association of demographic variables, comorbidities and symptoms with risk of death, admission to intensive care unit (ICU) or high-dependency unit (HDU), and use of invasive mechanical ventilation (IMV). Individuals were censored if they were lost to follow-up (for example, transferred to another facility) or remained in hospital on 24 May 2021. Time from symptom onset to the time of death or censoring (time last known to be alive), whichever occurred earlier, was used as the time scale, with patients considered at risk from the time of symptom onset or admission, whichever occurred later. Censoring times of discharged patients were modified and set to be equal to the latest censoring time (to account for informative censoring). For associations with admission to ICU, the time scale was from symptom onset to the earliest of admission to ICU, death, discharge, and censoring. The event was admission to ICU, death, or discharge, whichever occurred earliest. For associations with receipt of IMV, the time scale was from symptom onset to the earliest of IMV, death, discharge, and censoring. The event was IMV, death, or discharge, whichever occurred earliest. Patients were considered at risk from the time of symptom onset or admission, whichever occurred later. Models were adjusted for age and sex and stratified by country. We grouped countries with fewer than 50 individuals into a single category. We assessed the proportional hazards assumption using scaled Schoenfeld residuals. For explanatory variables with multiple categories (such as age groups), we used quasi-standard errors (15) to facilitate comparisons between any two groups.

We repeated the main analyses for patients with laboratory-confirmed SARS-CoV-2 only, as a sensitivity analysis. Analysis was performed using R version 4.0.5 and packages *survival, ggplot2, qvcalc* and *finalfit*.

## Results

### Participants’ characteristics

439,922 patients (**Figure 1)** were recruited from 720 sites (**Supplementary Table 1**) in 49 countries (**Figure 2** and **Supplementary figure 1**). Overall, 91.7% of the participants included in the primary analysis had a positive SARS-CoV-2 PCR test (**Table 1**). 50.1% were male and the median age was 60 (range 0 to 119, interquartile range [IQR] 29). The median time from symptom onset to admission was 2 (IQR 7) days. The median time from the latest of symptom onset or admission to discharge, death or date last known to be alive was 6 (IQR 15) days. Among 431,783 individuals with a known ICU admission status, 16.3% were admitted to ICU, about a third of whom were admitted directly (on the day of hospitalisation). Oxygen saturation on presentation to hospital was reported for 41.7% of patients with a median SpO2 of 96% (IQR 4%).

**Figure 2:**
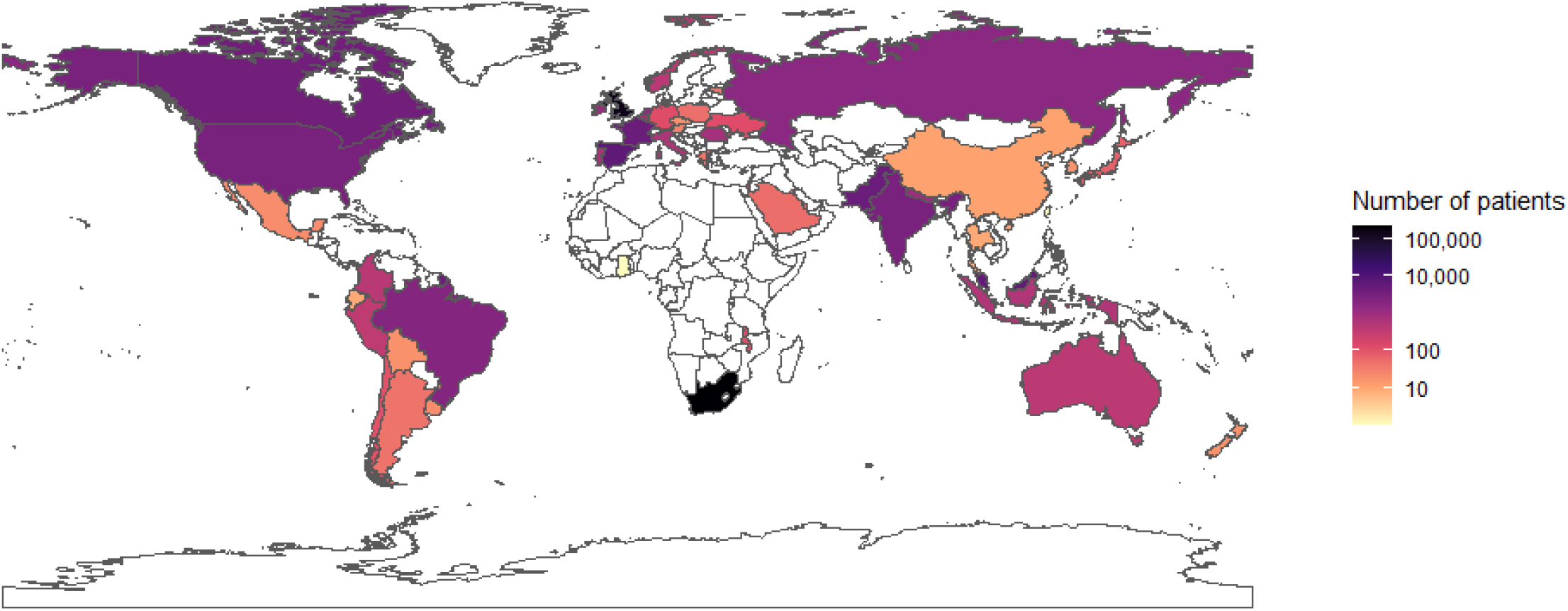
Numbers of patients by country.

**Table 1:** Participants’ characteristics. *Percent of individuals with oxygen saturation recorded

|  | N recorded (%) |  | Female | Male | Total |
| --- | --- | --- | --- | --- | --- |
| Total N |  |  | 219457 | 220465 | 439922 |
| Age | 439922 (100.0) | Median (IQR) | 59.0 (43.0 to 74.0) | 60.0 (47.0 to 74.0) | 60.0 (45.0 to 74.0) |
|  |  | 0-9 | 2365 (1.1) | 2906 (1.3) | 5271 (1.2) |
|  |  | 10-19 | 4114 (1.9) | 3195 (1.4) | 7309 (1.7) |
|  |  | 20-29 | 14203 (6.5) | 8479 (3.8) | 22682 (5.2) |
|  |  | 30-39 | 24994 (11.4) | 18779 (8.5) | 43773 (10.0) |
|  |  | 40-49 | 27858 (12.7) | 29770 (13.5) | 57628 (13.1) |
|  |  | 50-59 | 39141 (17.8) | 42737 (19.4) | 81878 (18.6) |
|  |  | 60-69 | 36659 (16.7) | 42362 (19.2) | 79021 (18.0) |
|  |  | 70-79 | 31832 (14.5) | 37701 (17.1) | 69533 (15.8) |
|  |  | 80-89 | 27853 (12.7) | 27417 (12.4) | 55270 (12.6) |
|  |  | 90+ | 10438 (4.8) | 7119 (3.2) | 17557 (4.0) |
| Region | 439922 (100.0) | East Asia & Pacific | 2265 (1.0) | 5205 (2.4) | 7470 (1.7) |
|  |  | Europe & Central Asia | 81043 (36.9) | 100488 (45.6) | 181531 (41.3) |
|  |  | Latin America & Caribbean | 1232 (0.6) | 1958 (0.9) | 3190 (0.7) |
|  |  | Middle East & North Africa | 201 (0.1) | 306 (0.1) | 507 (0.1) |
|  |  | North America | 2929 (1.3) | 4119 (1.9) | 7048 (1.6) |
|  |  | South Asia | 2874 (1.3) | 6181 (2.8) | 9055 (2.1) |
|  |  | Sub-Saharan Africa | 128913 (58.7) | 102208 (46.4) | 231121 (52.5) |
| Onset to admission (days) | 258289 (58.7) | Median (IQR) | 2.0 (0.0 to 7.0) | 3.0 (0.0 to 7.0) | 2.0 (0.0 to 7.0) |
| Length of hospital stay (days) | 439922 (100.0) | Median (IQR) | 5.0 (0.0 to 15.0) | 6.0 (1.0 to 16.0) | 6.0 (1.0 to 16.0) |
| Body mass index (BMI) (kg/m²) | 13949 (3.2) | Median (IQR) | 28.1 (23.8 to 33.0) | 27.5 (24.4 to 31.2) | 27.7 (24.2 to 31.8) |
| Heart rate on admission (bpm) | 174714 (39.7) | Median (IQR) | 89.0 (78.0 to 102.0) | 90.0 (78.0 to 103.0) | 90.0 (78.0 to 103.0) |
| Systolic blood pressure on admission (mmHg) | 181293 (41.2) | Median (IQR) | 129.0 (115.0 to 145.0) | 130.0 (116.0 to 144.0) | 130.0 (116.0 to 144.0) |
| Diastolic blood pressure on admission (mmHg) | 181361 (41.2) | Median (IQR) | 73.0 (65.0 to 82.0) | 75.0 (67.0 to 84.0) | 74.0 (66.0 to 83.0) |
| Temperature on admission (degrees C) | 182514 (41.5) | Median (IQR) | 37.0 (36.5 to 37.7) | 37.0 (36.5 to 37.9) | 37.0 (36.5 to 37.8) |
| Oxygen saturation on admission (%) | 183304 (41.7) | Median (IQR) | 96.0 (93.0 to 98.0) | 95.0 (93.0 to 97.0) | 96.0 (93.0 to 97.0) |
| - On oxygen therapy | 57687 (31.5% *) | Median (IQR) | 95.0 (92.0 to 97.0) | 95.0 (92.0 to 97.0) | 95.0 (92.0 to 97.0) |
| - In room air | 116762 (63.7% *) | Median (IQR) | 96.0 (94.0 to 98.0) | 96.0 (93.0 to 98.0) | 96.0 (93.0 to 98.0) |
| - Unknown oxygenation status | 9042 (4.9% *) | Median (IQR) | 96.0 (93.0 to 98.0) | 96.0 (93.0 to 97.0) | 96.0 (93.0 to 98.0) |
| Respiratory rate on admission | 175080 (39.8) | Median (IQR) | 20.0 (18.0 to 24.0) | 20.0 (18.0 to 26.0) | 20.0 (18.0 to 25.0) |
| Admission to ICU | 417879 (95.0) | Never | 187641 (89.8) | 173973 (83.3) | 361614 (86.5) |
|  |  | Later admission | 14037 (6.7) | 20338 (9.7) | 34375 (8.2) |
|  |  | Direct admission | 7251 (3.5) | 14639 (7.0) | 21890 (5.2) |
| Outcome | 439922 (100.0) | Unknown outcome | 13833 (6.3) | 16948 (7.7) | 30781 (7.0) |
|  |  | Death | 46498 (21.2) | 55683 (25.3) | 102181 (23.2) |
|  |  | Discharge | 159126 (72.5) | 147834 (67.1) | 306960 (69.8) |
| SARS-CoV-2 status | 439922 (100.0) | Positive | 198671 (90.5) | 204761 (92.9) | 403432 (91.7) |
|  |  | Unknown | 20786 (9.5) | 15704 (7.1) | 36490 (8.3) |

The majority of patients were recruited in two countries (South Africa [52.5%] and United Kingdom [36.7%]) (**Table 1**). Participants’ characteristics varied between patients admitted directly to ICU, those admitted at a later time point, and those never admitted (**Supplementary Table 1**). Anthropometric variables and vital signs varied by age (**Supplementary Table 2**).

### Presenting symptoms

The most common symptoms on presentation were fever, cough, and shortness of breath (**Figure 3** and **Supplementary Table 3**). There was some variation by country (**Supplementary figure 2**). Fatigue/malaise, cough and shortness of breath were most prevalent amongst patients 40 to 70 years old. The prevalence of altered consciousness/confusion increased with age and was reported in 30.2% of patients over 80 years of age. Loss or altered smell or taste were not commonly reported, with a high proportion of missing values for these two symptoms (39.1% for loss of smell and 40.5% for taste). We have previously described the associations of age and gender with presenting symptoms (16). Prevalence of symptoms by age was similar when we restricted our analysis to patients with laboratory-confirmed SARS-CoV-2 (**Supplementary figure 3**), but there were more missing values (**Supplementary figure 4**). Altered consciousness/confusion, cough, fatigue/malaise, fever, shortness of breath and vomiting/nausea were more frequently reported in patients with laboratory-confirmed SARS-CoV-2 infection than in those with a clinical diagnosis alone (**Supplementary figure 5**).

**Figure 3:**
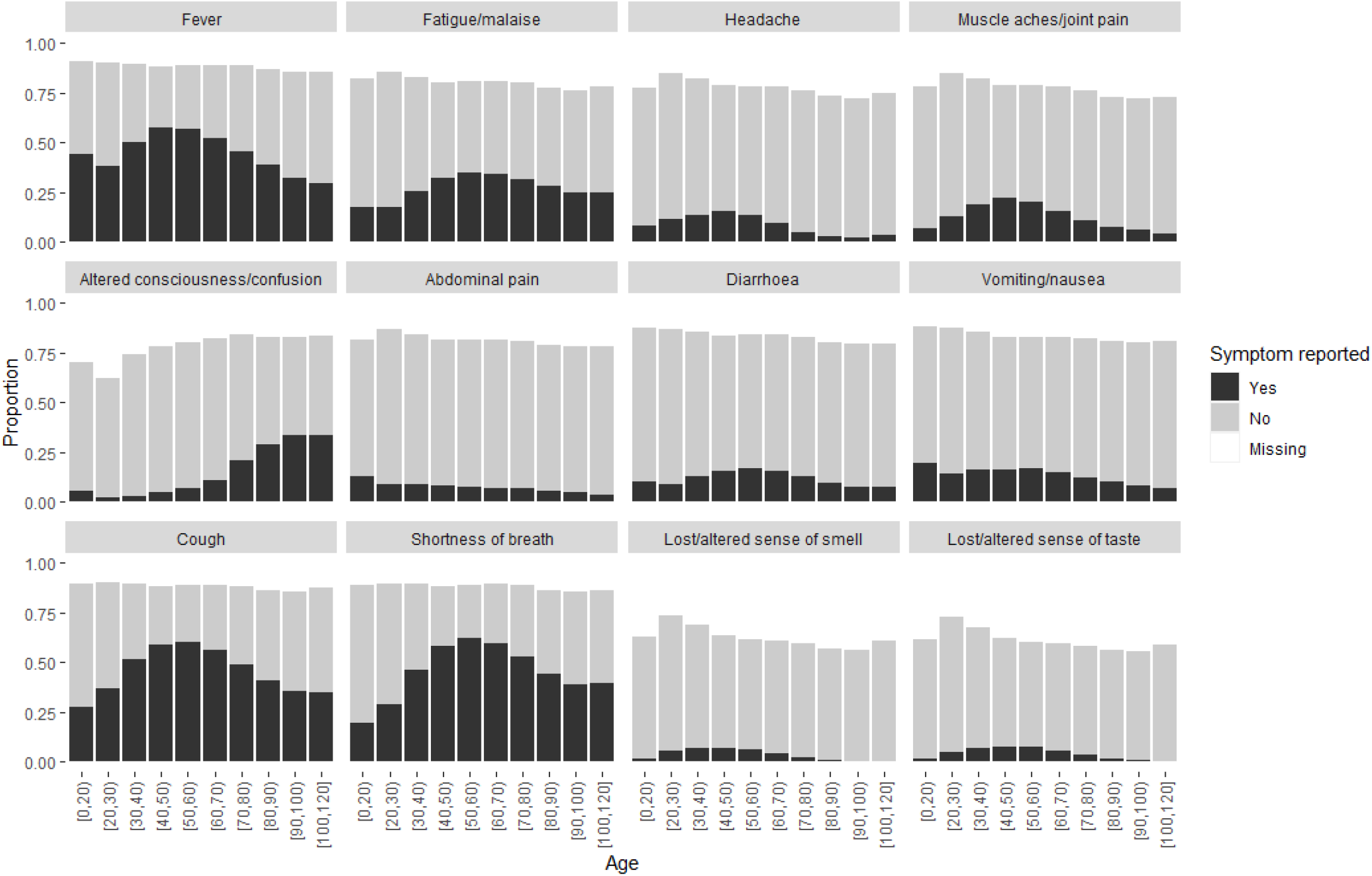
Symptom prevalence by age.

Overall, 50-75% of patients met one of the international symptom-based case definitions. This proportion was higher among those with laboratory-confirmed SARS-CoV-2 infection (**Supplementary figure 6**). Individuals aged 40-70 years were more likely to meet one of the four case definitions based on symptoms than patients at the extremes of the age distribution (**Figure 4**). Adults with laboratory-confirmed SARS-CoV-2 infection were more likely to meet one of the four case definitions based on symptoms than those with only a clinical diagnosis of SARS-CoV-2 infection, but the opposite was true amongst patients under 20 (**Supplementary figure 7**).

**Figure 4:**
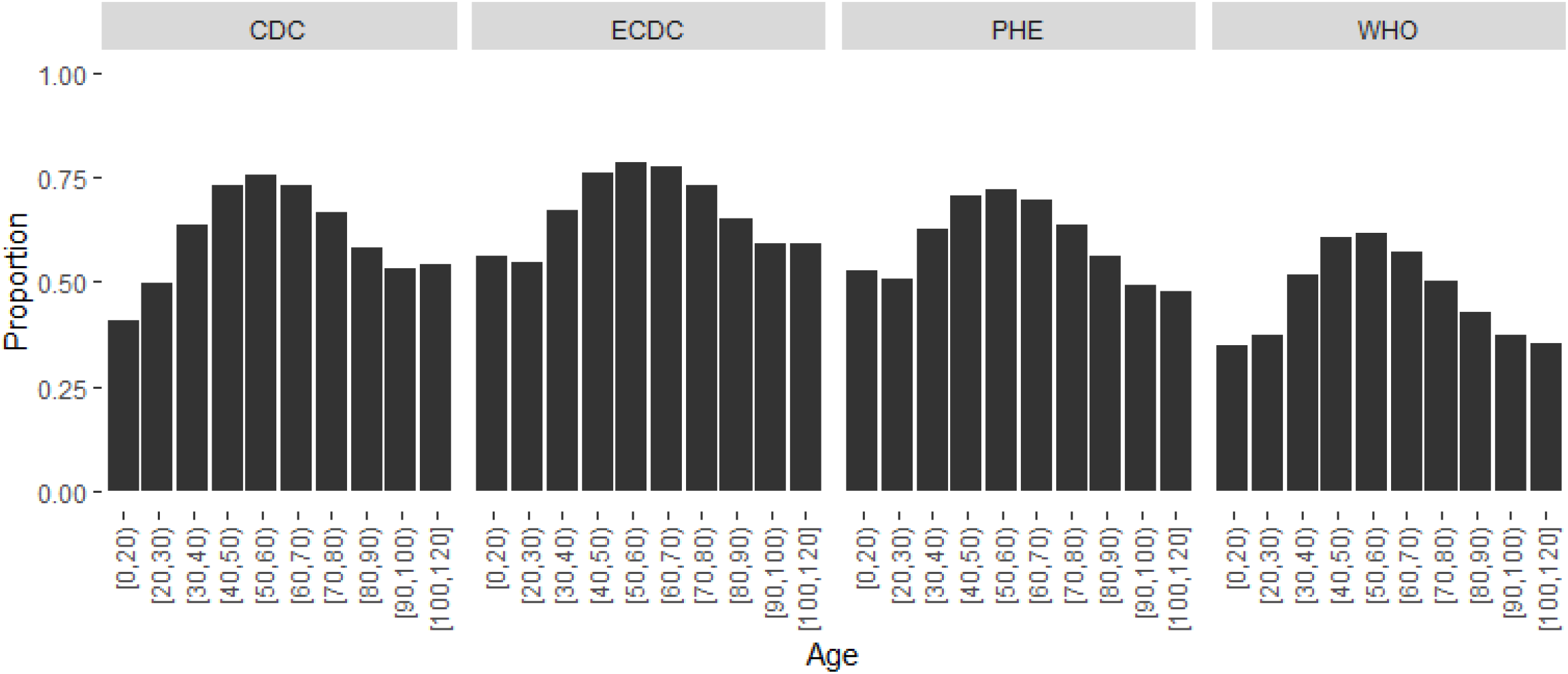
Proportions meeting each symptom definition by age.

Routine blood test results are shown in **Supplementary Table 4**. Median lymphocyte count was low (0.9 10^9^;IQR 0.6-1.3 10 cells per litre) among participants with available data, but median liver transaminase levels and median urea level were unremarkable. Several median values for routine blood test results varied with age; with increasing age, median lymphocyte count decreased, whereas median urea increased (**Supplementary figure 8**).

### Pre-existing comorbidities and risk factors

The most common pre-existing comorbidities were hypertension, diabetes, and chronic cardiac disease (**Figure 5** and **Supplementary Table 5**). Among 364,119 individuals with data available for any five or more comorbidities or risk factors, 107,292 (29.5%) had no comorbidities reported. The prevalence of most comorbidities varied by age (**Supplementary figure 9**). The prevalence of chronic cardiac disease, chronic kidney disease, dementia, hypertension and rheumatologic disorder increased with age. The prevalence of diabetes was highest in individuals aged 60 to 80. There were 16,381 patients with HIV infection, 6,708 with tuberculosis and 3,024 with both. 15,254 of the patients with HIV infection and 6541 patients with tuberculosis were from South Africa.

**Figure 5:**
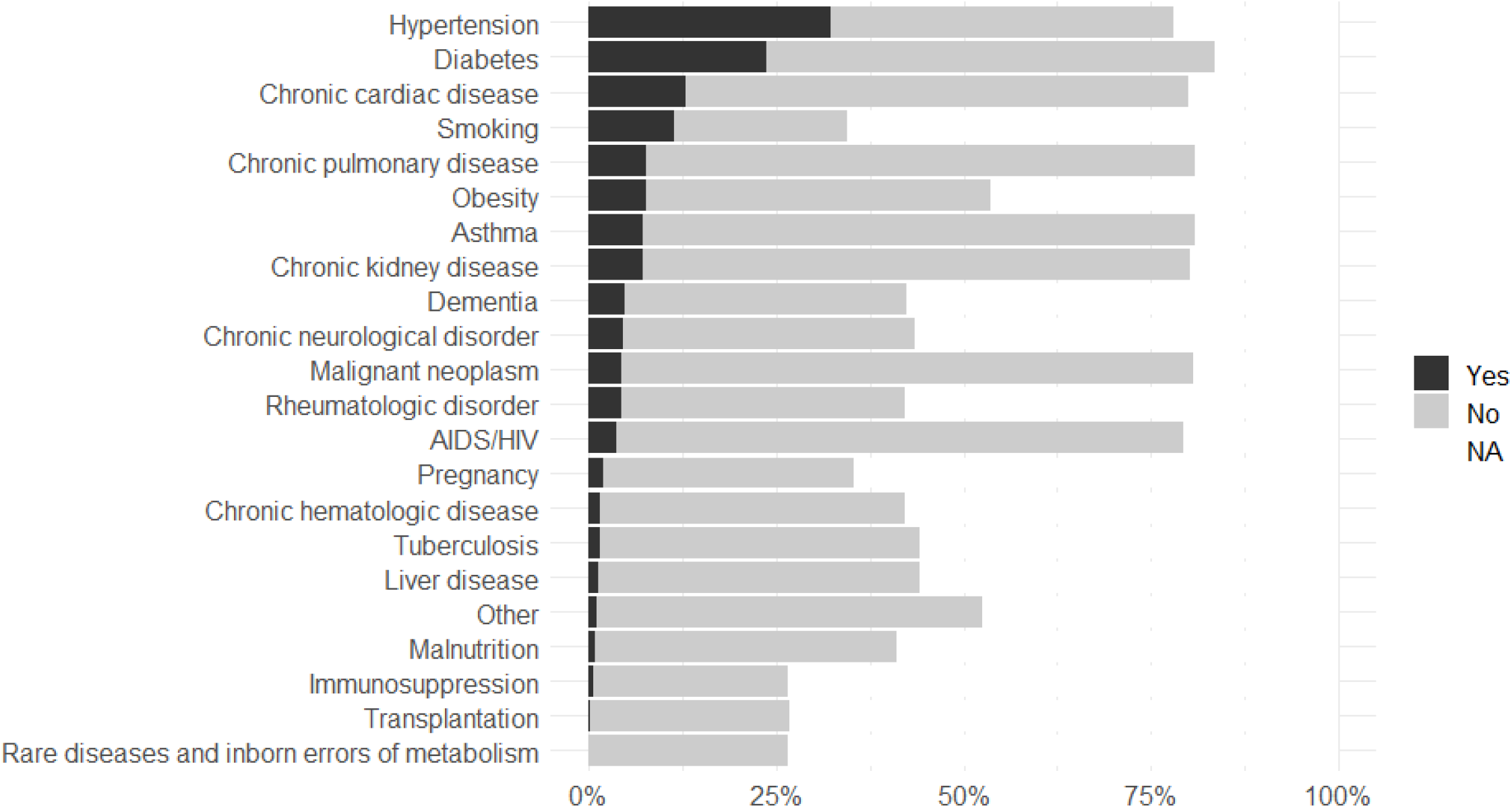
Prevalence of pre-existing comorbidities and risk factors.

### Treatments

Of 204,459 patients with available data on oxygen therapy (97.8% of total), 132,461 (64.8%) received oxygen therapy, which was delivered via high-flow nasal cannula to 40,441 (19.4%), by NIV to 31,132 (14.9%) and IMV to 25,362 (12.1%) (**Table 2**). As might be expected, a proportion of patients received multiple types of oxygen delivery systems during their admission (25,027 [18.9%]). For instance, 43% of those receiving IMV also received oxygen delivered via NIV.

**Table 2:** Oxygen supplementation. Among patients who received at least one type of oxygen supplementation

|  | N recorded (%) |  | Female | Male | Total |
| --- | --- | --- | --- | --- | --- |
| Total N (%) |  |  | 53664 (40.5) | 78797 (59.5) | 132461 |
| Extracorporeal membrane oxygenation | 128694 (97.2) | No | 51847 (99.2) | 75543 (98.8) | 127390 (99.0) |
|  |  | Yes | 406 (0.8) | 898 (1.2) | 1304 (1.0) |
| Invasive mechanical ventilation | 129708 (97.9) | No | 44676 (85.0) | 59670 (77.4) | 104346 (80.4) |
|  |  | Yes | 7901 (15.0) | 17461 (22.6) | 25362 (19.6) |
| Non-invasive ventilation | 131229 (99.1) | No | 42400 (79.7) | 57697 (73.9) | 100097 (76.3) |
|  |  | Yes | 10768 (20.3) | 20364 (26.1) | 31132 (23.7) |
| High-flow nasal cannula | 118809 (89.7) | No | 32886 (68.3) | 45482 (64.4) | 78368 (66.0) |
|  |  | Yes | 15246 (31.7) | 25195 (35.6) | 40441 (34.0) |
| Other or unspecified type of oxygen supplementation | 65979 (49.8) | Yes | 29632 | 36347 | 65979 |
| Number of types of oxygen supplementation* | 66482 (50.2) | 1 | 15763 (65.6) | 25692 (60.5) | 41455 (62.4) |
|  |  | 2 | 6322 (26.3) | 12212 (28.8) | 18534 (27.9) |
|  |  | 3 | 1874 (7.8) | 4382 (10.3) | 6256 (9.4) |
|  |  | 4 | 73 (0.3) | 164 (0.4) | 237 (0.4) |

\* ECMO, IMV, NIV or high-flow nasal cannula

The most used treatments were oxygen therapy, antibacterial agents and corticosteroids (**Figure 6** and **Supplementary Table 6**). The proportion of patients receiving antibacterial agents increased with age, as did the proportion receiving corticosteroids up to ages 70-80 (**Supplementary figure 10**). Information on antibacterial treatment was available for 188,595 patients, 149,900 (79.5%) of whom received antibacterial agents. 82,808 of 194,033 of patients with data available (42.7%) received corticosteroids. The use of corticosteroids increased after the publication of results of the RECOVERY trial (17) in June 2020 (**Supplementary figure 11**).

**Figure 6:**
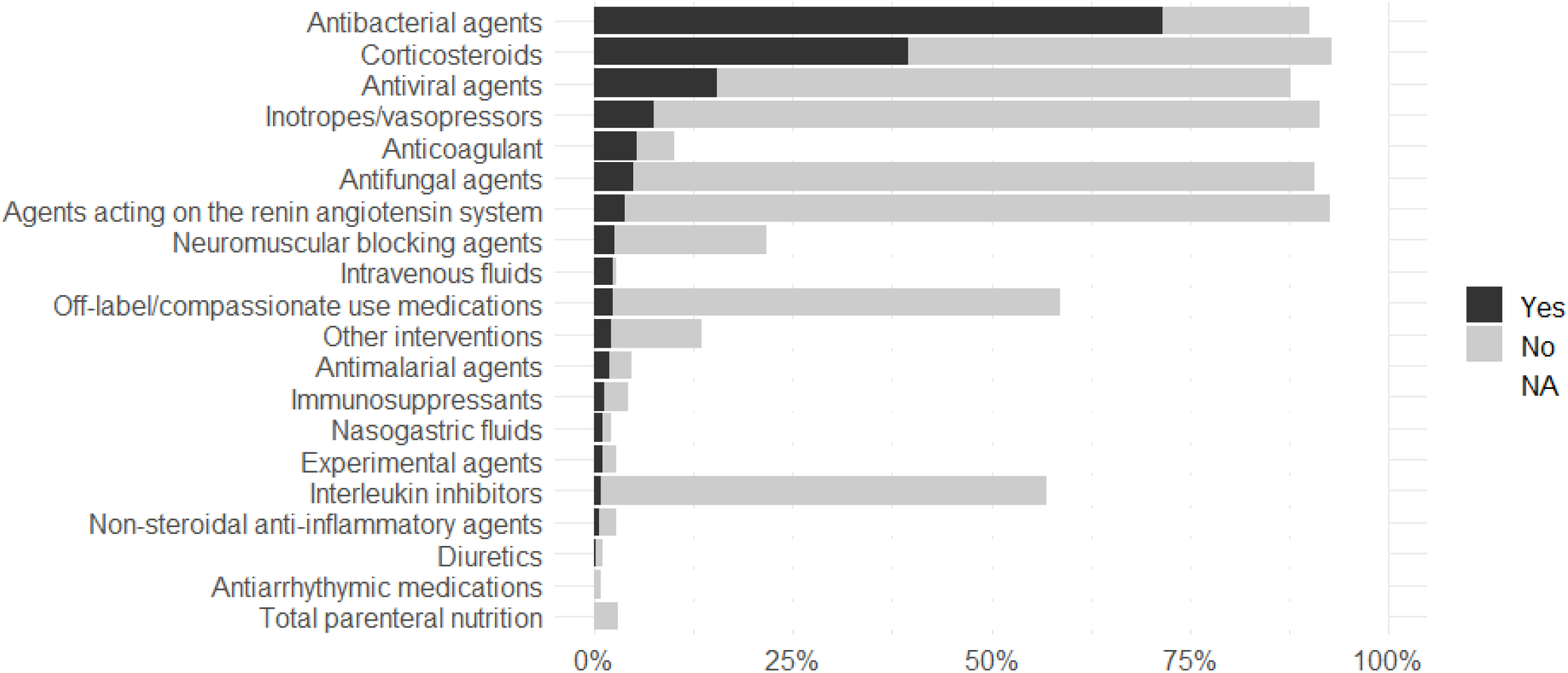
Proportion who have received each treatment.

### Case fatality ratio (CFR)

CFR varied by country (**Figure 7**). The weighted average CFR was 0.206 (SE 0.000522). Among patients for whom reporting commenced in the ICU, the CFR was 0.443 (SE 0.00349). Among patients admitted to the ICU but for whom reporting did not commence in the ICU, the CFR was 0.357 (SE 0.00254). The CFR varied over time during the study, as did patient recruitment at different sites (**Supplementary figure 12**). Admission criteria likely varied by country and time, contributing to the heterogeneity in illness severity. Death and discharge rates increased over the first 40 days since the latest of hospital admission and symptom onset (**Supplementary figure 13**).

**Figure 7:**
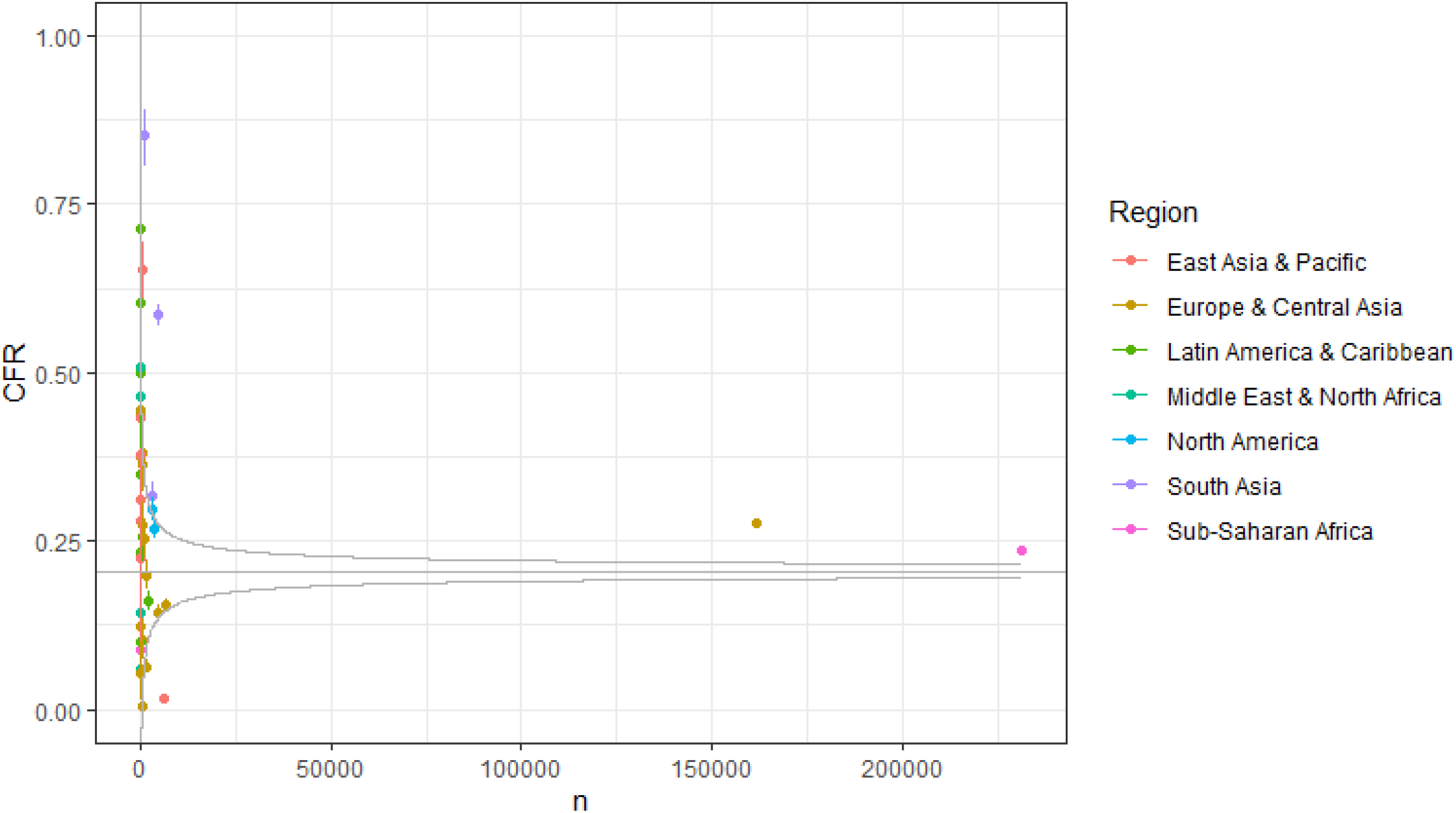
CFR by country. Each point is a country and points are coloured by region. Line is inverse variance weighted average CFR.

### Associations with death

The risk of death was higher for males than females (**Figure 8**). Older age was associated with a significantly higher risk of death, with a hazard ratio (HR) of 1.49 (95% CI 1.48, 1.50) per 10 years higher age, adjusting for sex and stratifying by country, with individuals aged 90-100 having an HR of 15.66 (95% CI 14.31, 17.13) compared to the 20-30 age group (**Supplementary figure 14**). Males had a significantly higher risk of death than females, with an HR of 1.26 (95% CI 1.24, 1.28), adjusting for age (in 10-year groups) and stratifying by country. There was evidence of deviation from the proportional hazards assumption for both variables. There was no particular trend for age, but the magnitude of the association of male sex with death appeared to increase with increasing time from admission (or symptom onset for patients who developed symptoms after admission). A model stratified by sex was fitted to estimate associations of age with death taking this time-varying association of sex into account. HRs for age estimated by the two models were very similar. HRs were also estimated by sex and age, stratifying (allowing different baseline hazards) by country (**Figure 9**), by fitting a model stratifying (allowing different baseline hazards) by country. HRs for age (**Supplementary figure 15**) and sex (**Supplementary figure 16**) varied by country.

**Figure 8:**
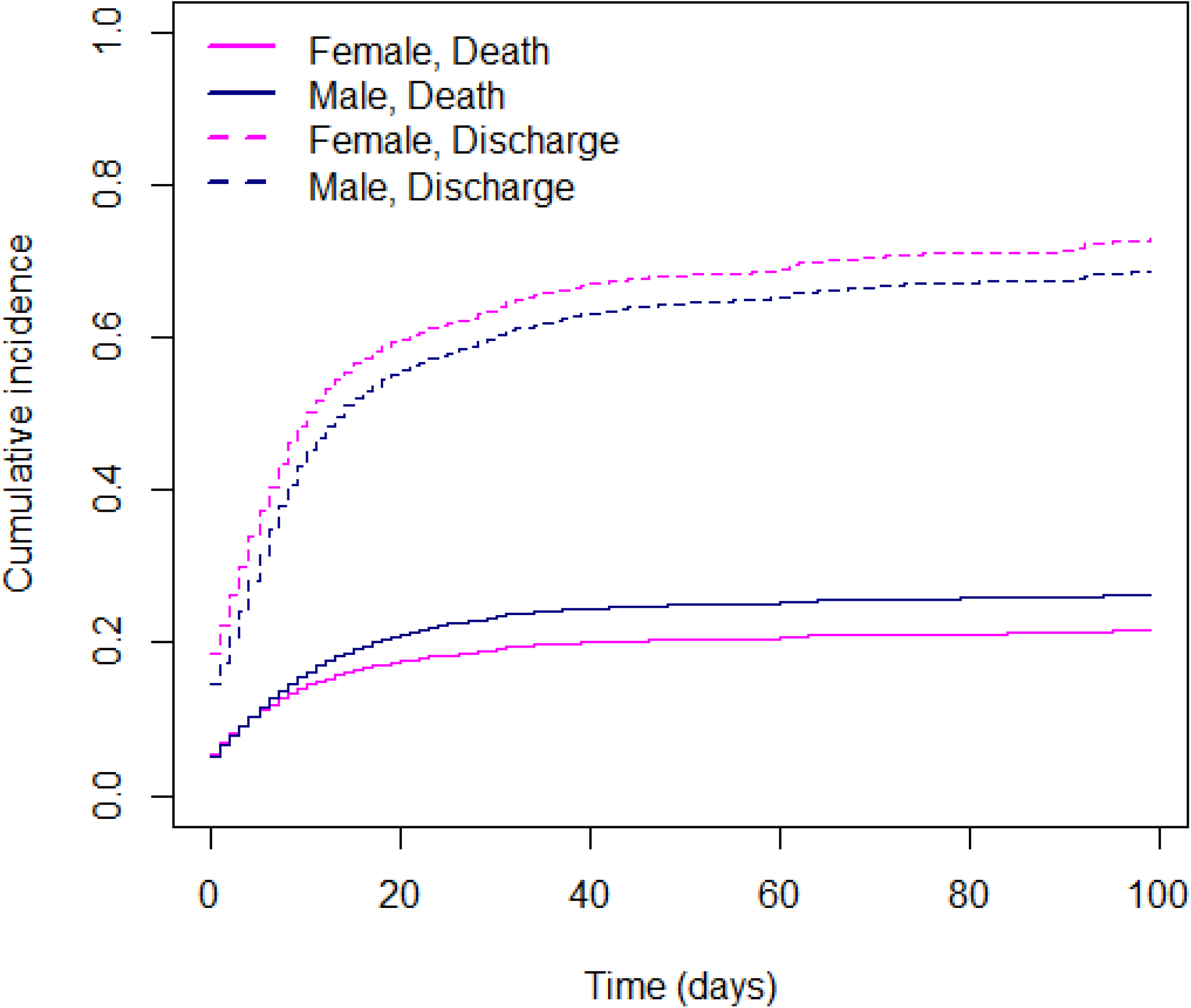
Cumulative incidence curves of death and discharge by sex.

**Figure 9:**
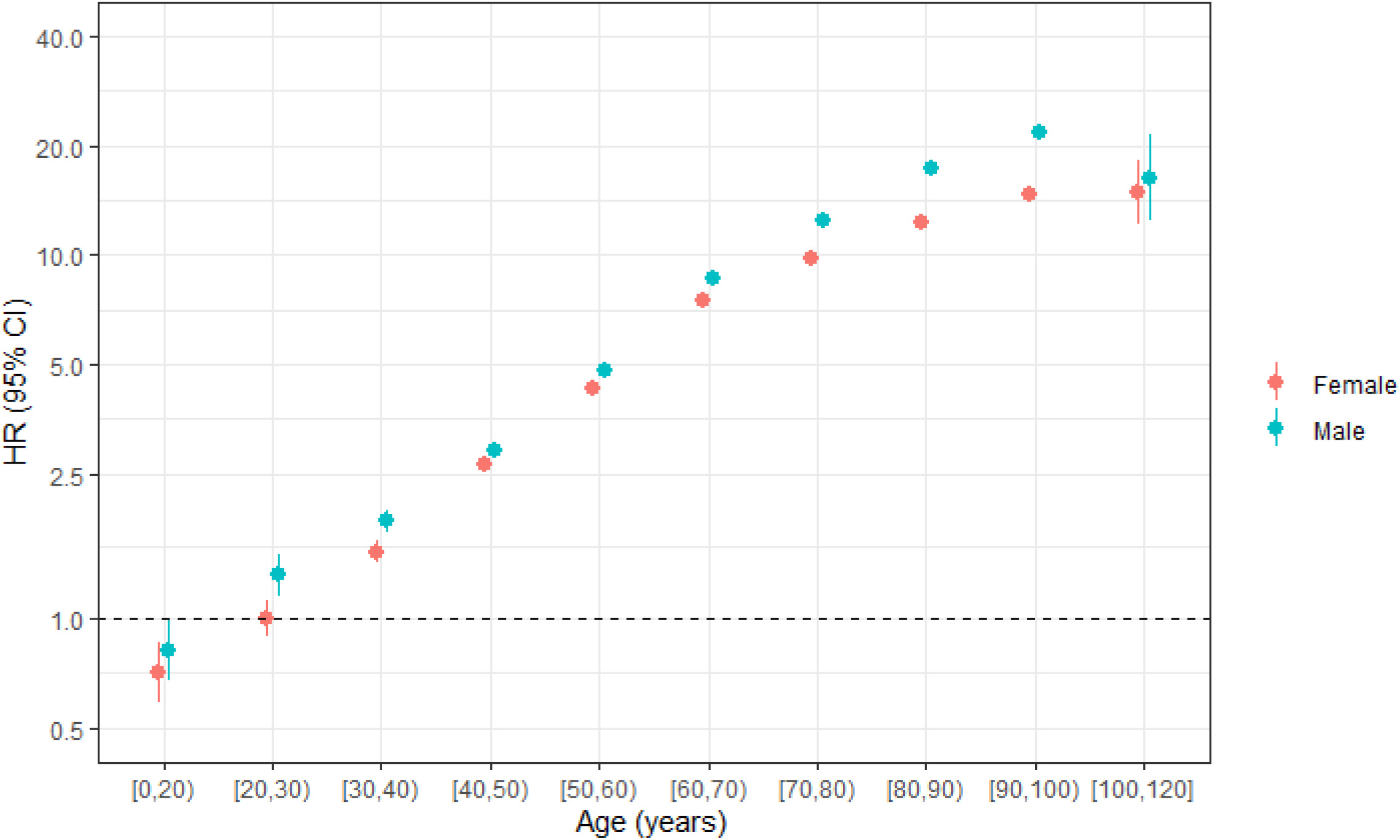
Hazard ratios and 95% CIs for death by age group and sex. Stratified by country

HRs of death were highest for patients with HIV infection (HR 1.87 [95% CI 1.78, 1.96]) and tuberculosis (1.86 [1.72, 2.02]), and there was some evidence of interaction (p = 0.05). Since many patients with these comorbidities were from South Africa, we assessed the associations separately among patients from South Africa and patients from all other countries by fitting a model including HIV, tuberculosis, age and age, stratified by sex and country. The associations with risk of death were similar between patients from South Africa and patients from other countries: respectively for HIV HR (95% CI) 1.60 (1.50, 1.71) and 1.26 (0.85, 1.86), and for tuberculosis 1.58 (1.43, 1.75) and 1.28 (0.87, 1.89). Transplantation (1.50 [1.32, 1.69]), immunosuppression (1.45 [1.35, 1.55]), chronic kidney disease (1.33 [1.30, 1.36]) and chronic pulmonary disease (1.32 [1.29, 1.35]) were associated with the highest relative risks of death. All reported comorbidities and risk factors were associated with a higher risk of death, except rheumatologic disorders for which there was no evidence of an association and pregnancy which was associated with a lower risk of death (HR 0.44 [0.38, 0.50] for females aged 15 to 45, adjusting for age, age^2^ and stratifying by country).

Shortness of breath (1.80 [1.76, 1.84]), inability to walk (1.76 [1.58, 1.96]), wheezing (1.45 [1.39, 1.50]), lymphadenopathy (1.43 [1.24, 1.63]), severe dehydration (1.37 [1.31, 1.42]), altered consciousness/confusion (1.31 [1.28, 1.34]), cough (1.22 [1.19, 1.24]), skin rash (1.22 [1.15, 1.30]), anorexia (1.19 [1.06, 1.34]), and fatigue/malaise (1.08 [1.06, 1.11]) were associated with a higher risk of death (**Figure 10**). In general, gastrointestinal, musculoskeletal symptoms, and loss of or altered taste or smell were associated with a lower risk of dying; for example nausea/vomiting had a HR of 0.83 [0.80, 0.86], abdominal pain 0.86 [0.83, 0.90], diarrhoea 0.94 [0.92, 0.97], and muscle aches/joint pain 0.89 (0.86, 0.92).

**Figure 10:**
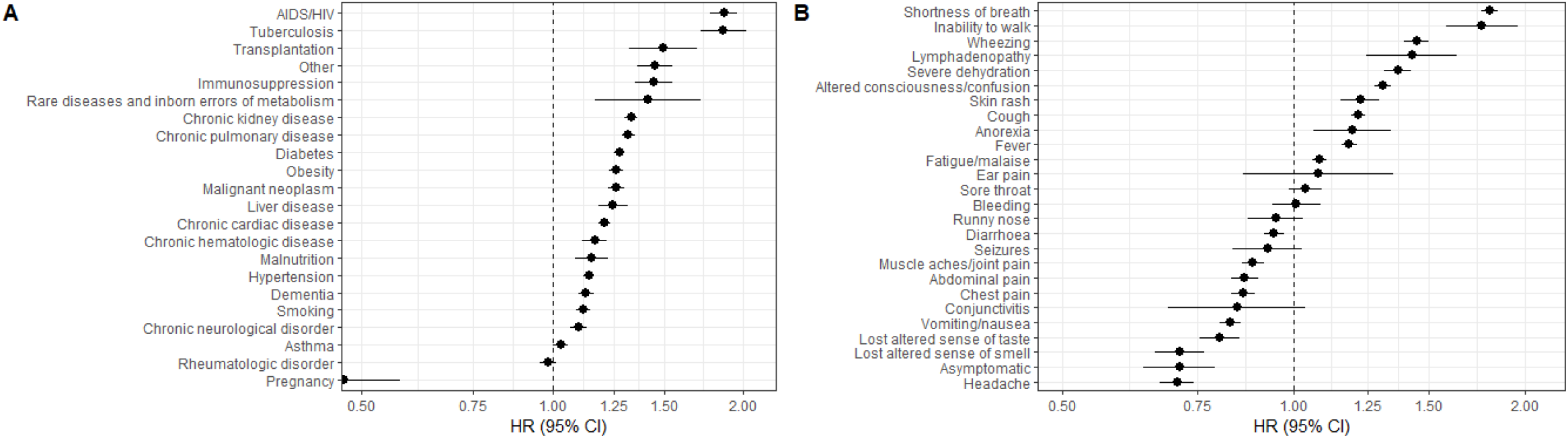
Associations of (A) comorbidities and (B) symptoms with risk of death. Adjusted for age and age^2^, stratified by sex and country

### Associations with admission to ICU and with use of IMV

The risk of admission to ICU increased with age after age 20-30 and started decreasing from age 60, with patients over 80 being very unlikely to be admitted to ICU. Men were more likely to be admitted to ICU overall, with a HR of 1.29 (1.27, 1.32). There was evidence of non-proportional hazards, indicating that the relative risk changed with time since symptom onset (or hospitalisation). There were similar patterns for risk of IMV (**Figure 11**).

**Figure 11:**
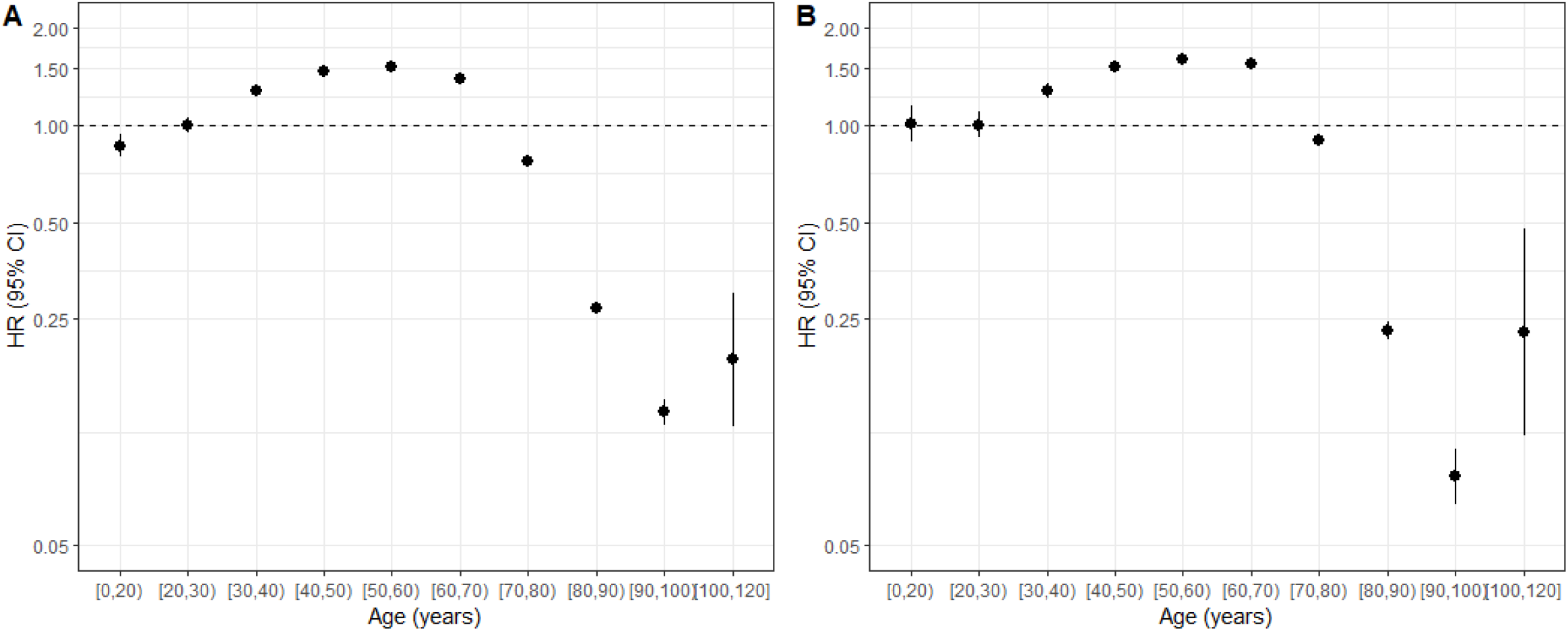
HRs (95% CIs) for (A) ICU admission and (B) use of IMV by age.

## Discussion

The ISARIC international cohort study includes standardized data on almost half a million patients from 720 sites across 49 countries. To our knowledge, this is the most extensive in-hospital COVID-19 cohort study in the world. The study’s size and breadth allow us to evaluate the contribution of individual risk factors to outcomes such as death, admission to intensive care, and use of mechanical ventilation. The value of the international cohort design is its capacity to cover the breadth of COVID-19 characteristics unencumbered by differences in classification and reporting. Furthermore, our international cohort design allowed us to explore risk factors that are globally uncommon, or uncommon in cohorts from high-income countries. For example, our dataset is the largest prospective cohort study of COVID-19 patients with HIV infection, tuberculosis, malnutrition, pregnancy, and transplantation.

Across the cohort, the most common presenting symptoms were fever, shortness of breath, and cough. Among other symptoms reported, the most common were altered consciousness in older patients and gastrointestinal symptoms in younger patients. Our data show that about one third of patients do not meet one of the four most widely used case definitions at the time of hospitalisation, particularly those in the younger and older age groups. These differences are relevant when defining testing or isolation and for early detection of new clusters and variants. Although case definitions must be simple, age-specific definitions may improve sensitivity. This has implications also for case management; about one third of patients did not require any oxygen therapy during their hospitalization.

Our study confirms that the strong association between age and risk of death from Covid-19 is a global phenomenon. The elderly are at a significantly higher risk of death from Covid-19. Every decade of life adds a 50% risk of dying, with those above 90 having a 15-fold higher risk than 20-30 year-olds. Although similar results were shown globally for non-hospitalised cases (18,19), our study reproduces these results globally and amongst hospitalised patients. Risk of death with increasing age differed between the sexes, with men having an increased risk of death around one-third higher than the corresponding female ten-year age group. Such age- and sex-specific CFRs with a global perspective are critical to understanding the global in-hospital burden of Covid-19. The pattern holds across lower and higher-income countries. Interestingly, non-respiratory presentations were associated with lower risk of death.

We found five comorbidities to be strongly associated with risk of death. The most substantial risk factor was HIV infection. There was a high proportion of people living with HIV (PLWH) in this cohort. Whilst retrospective health records analyses have been performed previously (20–22), this is the most extensive international cohort study of Covid-19 in PLWH. A recent cohort study performed in South Africa (23) demonstrated that PLWH had an adjusted odds ratio of death of 1.34, 95% CI 1.27–1.43. Our study reinforces these findings and confirms that it is an international phenomenon. Unfortunately, we have no further detail on how well-controlled HIV infection was or on the levels of immunocompromise for PLWH in our study. The second strongest association with the risk of death was a diagnosis of tuberculosis. To our knowledge, this is also the largest international cohort of patients co-infected with SARS-CoV-2 and tuberculosis (over 1,000 patients). There was also an interaction between HIV and tuberculosis. There have been few studies on the effect of COVID-19 on transplant patients. The 1067 transplant patients included in our dataset make it one of the largest cohorts to date. Overall, risk of death was about 50% higher in transplant patients, and 45% higher in those on immunosuppressive therapies. Our results suggest pregnancy is associated with a lower risk of death among people admitted to hospital, which appears to contrast with other studies suggesting an increased risk of death, intubation, or ICU admission for pregnant women (24). However, the UK Obstetric Surveillance System found that 55% of hospital admissions for pregnant women with COVID-19 were for the purpose of giving birth (25), whereas very few other elective and semi-elective admissions were taking place during the pandemic; this is likely to have increased the proportion of pregnant women in hospital with less severe Covid-19, compared with the broader cohort, confounding our observed lower risk of death for pregnant women.

Globally, case fatality ratios were much higher in the 5.2% of patients who were admitted to the ICU on the first day of their admission than those who required ICU at any point during their admission. The risk of admission to ICU increased with age, but then started decreasing from age 60 years, with patients over 80 years being very unlikely to be admitted to ICU. Compared with other studies, these results are consistent for patients below age 60 years, but not for those above 60 years. For example, in a study from the United States (26) and in a separate meta-analysis (27), elderly patients were more likely to be admitted to ICU than their younger counterparts (28); this may reflect geographical variation in clinical practice.

This international cohort study overcomes some of the traditional problems of observational studies by using standardised variables and outcome measures. Our data should be crucial for modelling and health system planning. For example, we note the greatly increased risk of death amongst patients with tuberculosis and malnutrition in our cohort and protecting such individuals from Covid-19 must be a critical public health priority for countries with high prevalence rates of these conditions. Equally concerning is our finding of increased risk of death amongst PLWH. Much of the world’s PLWH population resides in Sub-Saharan Africa; our data may indicate a phenomenon that is currently hidden due to under-testing for COVID-19 (29) across Africa.

While our study includes a broad range of data from different countries, various sites have different levels of data completeness. For example, we cannot evaluate the proportion of patients with HIV infection or tuberculosis who were taking appropriate, effective treatments. We have no further detail on the type of organ received by the transplantation cohort. While we have produced a ‘snapshot’ of the association of risk factors and outcomes in COVID-19, pandemics are complex, dynamic phenomena. Country-variations in disease incidence during different time points complicate comparisons. Our data will increasingly be influenced by the provision of vaccination and effective treatments, as well as the variability in access to these measures in the global context. We do not include data on SARS-CoV-2 variants of concern in this paper. The majority of submitted case records come from two countries, the United Kingdom and South Africa.

## Conclusion

This paper represents the largest international cohort of hospitalized COVID-19 patients published to date. We demonstrate several associations of global importance, including an increased risk of death in patients with HIV and TB. The ISARIC global collaboration continues to collect standardized data which will enable continued data-led comparisons as the world implements vaccination, treatment, and public health control strategies.

## Data sharing

The ISARIC-WHO Clinical Characterisation Protocol, case report form and consent forms are openly available on the ISARIC website at https://isaric.org/research/covid-19-clinical-research-resources/clinical-characterisation-protocol-ccp/. The statistical analysis plan is openly available on the ISARIC website at https://isaric.org/research/covid-19-clinical-research-resources/accessing-covid-19-clinical-data/approved-uses-of-platform-data/

Most individual patient data are available to researchers approved by the Data Access Committee. The data inventory, application form and terms of access for the COVID-19 Data Platform, hosted by the Infectious Diseases Data Observatory (IDDO), are available at https://www.iddo.org/covid19/data-sharing/accessing-data.

All individual participant data are available to individuals from sites who have contributed to the ISARIC COVID-19 Platform via the ISARIC Partner Analysis Scheme. See details via this link: https://isaric.org/research/isaric-partner-analysis-frequently-asked-questions/

## Data Availability

Data sharing
The ISARIC-WHO Clinical Characterisation Protocol, case report form and consent forms are openly available on the ISARIC website at https://isaric.org/research/covid-19-clinical-research-resources/clinical-characterisation-protocol-ccp/. The statistical analysis plan is openly available on the ISARIC website at https://isaric.org/research/covid-19-clinical-research-resources/accessing-covid-19-clinical-data/approved-uses-of-platform-data/
Most individual patient data are available to researchers approved by the Data Access Committee. The data inventory, application form and terms of access for the COVID-19 Data Platform, hosted by the Infectious Diseases Data Observatory (IDDO), are available at https://www.iddo.org/covid19/data-sharing/accessing-data.
All individual participant data are available to individuals from sites who have contributed to the ISARIC COVID-19 Platform via the ISARIC Partner Analysis Scheme. See details via this link: https://isaric.org/research/isaric-partner-analysis-frequently-asked-questions/
 

## Acknowledgments

This work was made possible by the UK Foreign, Commonwealth and Development Office; Wellcome [215091/Z/18/Z] [205228/Z/16/Z] [220757/Z/20/Z]; Bill & Melinda Gates Foundation [OPP1209135]; CIHR Coronavirus Rapid Research Funding Opportunity OV2170359 and the coordination in Canada by Sunnybrook Research Institute; endorsement of the Irish Critical Care-Clinical Trials Group, co-ordination in Ireland by the Irish Critical Care-Clinical Trials Network at University College Dublin and funding by the Health Research Board of Ireland [CTN-2014-12]; the Rapid European COVID-19 Emergency Response research (RECOVER) [H2020 project 101003589] and European Clinical Research Alliance on Infectious Diseases (ECRAID) [965313]; the COVID clinical management team, AIIMS, Rishikesh, India; Cambridge NIHR Biomedical Research Centre; the dedication and hard work of the Groote Schuur Hospital Covid ICU Team; support by the Groote Schuur nursing and University of Cape Town registrar bodies coordinated by the Division of Critical Care at the University of Cape Town; the Liverpool School of Tropical Medicine and the University of Oxford; the dedication and hard work of the Norwegian SARS-CoV-2 study team; the Research Council of Norway grant no 312780, and a philanthropic donation from Vivaldi Invest A/S owned by Jon Stephenson von Tetzchner; Imperial NIHR Biomedical Research Centre; Innovative Medicines Initiative Joint Undertaking under Grant Agreement No. 115523 COMBACTE, resources of which are composed of financial contribution from the European Union’s Seventh Framework Programme (FP7/2007-2013) and EFPIA companies, in-kind contribution; Stiftungsfonds zur Förderung der Bekämpfung der Tuberkulose und anderer Lungenkrankheiten of the City of Vienna, Project Number: APCOV22BGM; Italian Ministry of Health “Fondi Ricerca corrente– L1P6” to IRCCS Ospedale Sacro Cuore–Don Calabria; Australian Department of Health grant (3273191); Gender Equity Strategic Fund at University of Queensland, Artificial Intelligence for Pandemics (A14PAN) at University of Queensland, the Australian Research Council Centre of Excellence for Engineered Quantum Systems (EQUS, CE170100009), the Prince Charles Hospital Foundation, Australia; and preparedness work conducted by the Short Period Incidence Study of Severe Acute Respiratory Infection.

This work uses Data / Material provided by patients and collected by the NHS as part of their care and support #DataSavesLives. The Data / Material used for this research were obtained from ISARIC4C. The COVID-19 Clinical Information Network (CO-CIN) data was collated by ISARIC4C Investigators. Data and Material provision was supported by grants from: the National Institute for Health Research (NIHR; award CO-CIN-01), the Medical Research Council (MRC; grant MC_PC_19059), and by the NIHR Health Protection Research Unit (HPRU) in Emerging and Zoonotic Infections at University of Liverpool in partnership with Public Health England (PHE), (award 200907), NIHR HPRU in Respiratory Infections at Imperial College London with PHE (award 200927), Liverpool Experimental Cancer Medicine Centre (grant C18616/A25153), NIHR Biomedical Research Centre at Imperial College London (award IS-BRC-1215-20013), and NIHR Clinical Research Network providing infrastructure support.

## Potential Conflict of interest

Allavena, C. declares personal fees from ViiVHealthcare, MSD, Janssen and Gilead, all outside the submitted work.

Andréjak, C. declares personal fees for lecture from Astra Zeneca, outside the submitted work.

Borie, R. declares personal fees for lecture from Roche, Sanofi and Boehringer Ingelheim, outside the submitted work

Bosse, Hans Martin is co-investigator for placebo studies in infants and children in clinical trials by Actelion/Janssen (Johnson&Johnson), outside the submitted work

Cheng, M. declares grants from McGill Interdisciplinary Initiative in Infection and Immunity, grants from Canadian Institutes of Health Research, during the conduct of the study; personal fees from GEn1E Lifesciences (as a member of the scientific advisory board), personal fees from nplex biosciences (as a member of the scientific advisory board), outside the submitted work. He is the co-founder of Kanvas Biosciences and owns equity in the company. In addition, M. Cheng reports a patent Methods for detecting tissue damage, graft versus host disease, and infections using cell-free DNA profiling pending, and a patent Methods for assessing the severity and progression of SARS-CoV-2 infections using cell-free DNA pending.

Cholley, B. declares personal fees (for lectures and participation to advisory boards) from Edwards, Amomed, Nordic Pharma, and Orion Pharma.

Claure-Del Granado, R. declares personal fees (for lectures and participation to advisory boards) from Nova Biomedical, Medtronic, and Baxter all outside the submitted work.

Cruz-Bermúdez, J.L. declares personal fees from Elsevier for advice, outside the submitted work.

Cummings, M. and O’Donnell, M. participated as investigators for clinical trials evaluating the efficacy and safety of remdesivir (sponsored by Gilead Sciences) and convalescent plasma (sponsored by Amazon) in hospitalized patients with COVID-19. Support for this work is paid to Columbia University.

Dalton, H. declares personal fees for medical director of Innovative ECMO Concepts and honorarium from Abiomed/BREETHE Oxi-1 and Instrumentation Labs. Consultant fee, Entegrion Inc.

Deplanque, D. declares personal fees from Biocodex, Bristol-Myers Squibb and Pfizer (advisory boards)

Donnelly, C.A. declares research funding from the UK Medical Research Council and the UK National Institute for Health Research. Dudman, S. reports grants from Research Council of Norway grant no 312780.

Durante-Mangoni, E. declares funding via his Institution from MSD, Pfizer, and personal fees or participation in advisory boards or participation to the speaker’s bureau of Roche, Pfizer, MSD, Angelini, Correvio, Nordic Pharma, Bio-Merieux, Abbvie, Sanofi-Aventis, Medtronic, Tyrx and DiaSorin.

Dyrhol-Riise, AM, declares grants from Gilead outside this work.

Grasselli, G. declares personal fees from Getinge, Biotest, Draeger Medical, Fisher & Paykel, MSD and unrestricted research grant from MSD and Fisher & Paykel, all outside the submitted work.

Guerguerian AM. Participated as site investigator for the Hospital For Sick Children, Toronto, Canada as a site through SPRINT-SARI Study via the Canadian Critical Care Trials Group sponsored in part by the Canadian Institutes of Health Research.

Ho, A. declares grant funding from Medical Research Council UK, Scottish Funding Council - Grand Challenges Research Fund, and the Wellcome Trust, outside this submitted work.

Holter, J. C. reports grants from Research Council of Norway grant no 312780, and from Vivaldi Invest A/S owned by Jon Stephenson von Tetzchner, during the conduct of the study.

Kimmoun, A. declares personal fees (payment for lectures) from Baxter, Aguettant, Aspen.

Kumar, D. declares grants and personal fees from Roche, GSK and Merck; and personal fees from Pfizer and Sanofi.

Kutsogiannis, D.J. declares personal fees for a lecture from Tabuk Pharmaceuticals and the Saudi Critical Care Society

Kutsyna, G. declares the study consulting fee for clinical trial ClinicalTrials.gov Identifier: NCT04762628

Laffey, J. reports that he has received fees for consultancy from GlaxoSmithKline and from Baxter Therapeutics for work outside the scope of this work.

Lairez, O. declares grant funding from Pfizer; conference fees from Amicus, GE Healthcare, Novartis, Sanofi-Genzyme, and Takeda-Shire; and consultancy fees from Alnylam, Amicus, Pfizer, Takeda-Shire.

Lee, J. reports grants from European Commission PREPARE grant agreement No 602525, European Commission RECOVER Grant Agreement No 101003589 and European Commission ECRAID-Plan Grant Agreement 825715 supporting the conduct, coordination and management of the work.

Lee, T.C. declares research salary support from les Fonds de recherche du Québec – Santé.

Lefèvre, B. declares travel/accommodation/meeting expenses from Mylan and Gilead, all outside the submitted work.

Lellouche, F. declares grants from CIHR for COVID-19 studies, is co-founder and administrator of Oxynov.inc, fees from Fisher&Paykel, Vygon and Novus

Lemaignen, A. declares personal fees (payment for lectures) from MSD and Gilead; and travel/accommodation/meeting expenses from Pfizer.

Leone, M declares personal fees from Gilead, MSD, Aspen, Ambu and Amomed

Lescure, F.X. declares personal fees (payment for lectures) from Gilead, MSD; and travel/accommodation/meeting expenses from Astellas, Eumedica, MSD.

Liu, K. reports personal fees from MERA and receives a salary from TXP Medical completely outside the submitted work.

Martin-Blondel G declares support for attending meetings and personal fees from BMS, MSD, Janssen, Sanofi, Pfizer and Gilead for lectures outside the submitted work.

Martin-Loeches I. declared lectures for Gilead, Thermofisher, Pfizer, MSD; advisory board participation for Fresenius Kabi, Advanz Pharma, Gilead, Accelerate, Merck; and consulting fees for Gilead outside of the submitted work.

Martín-Quiros, A. declares consulting fees for Gilead.

Mentré F, declares consulting fees from IPSEN, Servier and Da Volterra, and reports research grants to her group from Sanofi, Roche, Servier and Da Voleterra, all outside the submitted work.

Montrucchio, G declares personal fees for lecture from Pfizer, Gilead outside the submitted work.

Murthy, S declares receiving salary support from the Health Research Foundation and Innovative Medicines Canada Chair in Pandemic Preparedness Research.

Nichol, A. declares a grant from the Health Research Board of Ireland to support data collection in Ireland (CTN-2014-012).

Openshaw, P. has served on scientific advisory boards for Janssen/J&J, Oxford Immunotech Ltd, GSK, Nestle and Pfizer (fees to Imperial College). He is Imperial College lead investigator on EMINENT, a consortium funded by the MRC and GSK. He is a member of the RSV Consortium in Europe (RESCEU) and Inno4Vac, Innovative Medicines Initiatives (IMI) from the European Union.

Peltan, I.D. declares grant support from the National Institutes of Health and, outside the submitted work, grant support from Centers for Disease Control and Prevention, and Jannsen and payments to his institution for trials by Regeneron and Asahi Kasei Pharma on which he served as an investigator.

Pesenti, A. declares personal fees from Maquet, Novalung/Xenios, Baxter, and Boehringer Ingelheim.

Peytavin G., declares consulting fees (for lectures and/or participation in advisory boards) and travel grants from Gilead Sciences, Janssen, Merck, Takeda, Theratechnologies, and ViiV Healthcare.

Poissy, J. declares personal fees from Gilead for lectures, outside the submitting work

Povoa, P. declares personal fees (for lectures and advisory boards) from MSD, Technophage, Sanofi, and Gilead.

Póvoas, D. declares consulting fees (for lectures and/or participation in advisory boards) from Roche and Viiv Healthcare; and travel/accommodation/meeting expenses from Abbvie, Gilead Sciences, Janssen Cilag, Merck Sharp & Dohme and ViiV Healthcare

Rewa, O. declares consulting fees from Baxter Inc.

Rossanese, A. declares consulting fees (for lectures and/or participation to advisory boards) from Emergent BioSolutions and Sanofi Pasteur, but all outside of the frame of the submitted work.

Săndulescu, O. has been an investigator in COVID-19 clinical trials by Algernon Pharmaceuticals, Atea Pharmaceuticals, Regeneron Pharmaceuticals, Diffusion Pharmaceuticals, and Celltrion, Inc., outside the scope of the submitted work.

Semple, M.G. reports grants from DHSC National Institute of Health Research UK, from the Medical Research Council UK, and from the Health Protection Research Unit in Emerging & Zoonotic Infections, University of Liverpool, supporting the conduct of the study; other interest in Integrum Scientific LLC, Greensboro, NC, USA, outside the submitted work.

Serpa Neto, A. declares personal lecture fees from Drager outside the submitted work.

Shrapnel, S. participated as an investigator for an observational study analysing ICU patients with COVID-19 (for the Critical Care Consortium including ECMOCARD) funded by The Prince Charles Hospital Foundation during the conduct of this study. S. Shrapnel reports in kind support from the Australian Research Council Centre of Excellence for Engineered Quantum Systems (CE170100009).

Streinu-Cercel, Adrian has been an investigator in COVID-19 clinical trials by Algernon Pharmaceuticals, Atea Pharmaceuticals, Regeneron Pharmaceuticals, Diffusion Pharmaceuticals, and Celltrion, Inc., outside the scope of the submitted work.

Streinu-Cercel, Anca has been an investigator in COVID-19 clinical trials by Algernon Pharmaceuticals, Atea Pharmaceuticals, Regeneron Pharmaceuticals, Diffusion Pharmaceuticals, and Celltrion, Inc., outside the scope of the submitted work.

Summers, C. reports that she has received fees for consultancy for Abbvie relating to COVID-19 therapeutics. She was also the UK Chief Investigator of a GlaxoSmithKline plc sponsored study of a therapy for COVID, and is a member of the UK COVID Therapeutic Advisory Panel (UK-CTAP). Outside the scope of this work, Dr Summers’ institution receives research grants from the Wellcome Trust, UKRI/MRC, National Institute for Health Research (NIHR), GlaxoSmithKline and AstraZeneca to support research in her laboratory.

Tedder, R. reports grants from MRC/UKRI during the conduct of the study. In addition, R. Tedder has a patent United Kingdom Patent Application No. 2014047.1 “SARS-CoV-2 antibody detection assay” issued.

Terzi, N. reports personal fees from Pfizer, outside the submitted work. JF Timsit participated in an advisory board for MSD, Pfizer, nabriva, Gilead, Shionoghi, Medimune outside the submitted work; personal fees from Merck, Pfizer, Gilead, Beckton dickinson, Medimune, Bayer pharma; lecture fees from MSD, Biomerieux, Pfizer, Shionoghi; and research grants to his research unit from Pfizerpers, Merck, Thermofisher.

Turtle, L. reports grants from MRC/UKRI during the conduct of the study and fees from Eisai for delivering a lecture related to COVID-19 and cancer, paid to the University of Liverpool.

Ullrich, R. reports grant funding to his institution from Apeptico, APEIRON, Biotest, Bayer, CCORE and Philips, as well as personal fees from Biotest. He holds European patent EP15189777.4 “Blood purification device” and equity in CCORE Technology GesmbH, a medical device research and development company.

Visseaux B. declares personal fees from BioMérieux, Qiagen and Gilead and research grants from Qiagen, all outside the submitted work.

West, E. reports grant funding from the Firland Foundation, the US CDC, and the Bill and Melinda Gates Foundation for studies of COVID-19, and grant funding from the US NIH for studies of other respiratory infections.

## AUTHOR LIST

**ISARIC Clinical Characterisation Group**

**CRediT author statement (based on Brand et al**., **2015, doi: 10.1087/20150211)**

**Conceptualization** (this analysis): Kartsonaki, Christiana; Olliaro, Piero L.; Merson, Laura; Dankwa, Emmanuelle A; Donnelly, Christl A.; Rojek, Amanda; Hall, Matthew; Semple, Malcolm G; Carson, Gail; Ho, Antonia Ying Wai; Sigfrid, Louise; Pritchard, Mark G. **Conceptualization** (study): Baillie, J. Kenneth; Begum, Husna; Burrell, Aidan; Carson, Gail; Dunning, Jake; Fowler, Robert A.; Horby, Peter; Marshall, John; McArthur, Colin; Merson, Laura; Nichol, Alistair; Parke, Rachael; Perren, Cobb J; Semple, Malcolm G.; Uyeki, Tim; Webb, Steve. **Methodology**: Kartsonaki, Christiana; Olliaro, Piero L.; Merson, Laura; Dankwa, Emmanuelle A; Donnelly, Christl A. **Software and formal analysis**: Kartsonaki, Christiana; Escher, Martina; Dankwa, Emmanuelle A; Hall, Matthew. **Data curation**: Citarella, Barbara Wanjiru; Kelly, Sadie; Kennon, Kalynn; Lee, James; Merson, Laura; Plotkin, Daniel; Smith, Sue; Strudwick, Samantha. **Administration**: Citarella, Barbara Wanjiru; Merson, Laura. **Writing - original draft**: Kartsonaki, Christiana; Olliaro, Piero L.; Dunning, Jake; Baruch, Joaquin; Dagens, Andrew; Sigfrid, Louise; Merson, Laura; Rojek, Amanda; Pritchard, Mark G.; Citarella, Barbara Wanjiru. **Visualization**: Kartsonaki, Christiana; Dankwa, Emmanuelle A; Baruch, Joaquin. **Writing - review and editing:** All authors. **Data Contributors-including Investigation, Supervision, Resources, Project administration and Funding acquisition:** Abbas, Ali; Abdukahil, Sheryl Ann; Abdulkadir, Nurul Najmee; Abe, Ryuzo; Abel, Laurent; Absil, Lara; Acharya, Subhash; Acker, Andrew; Adachi, Shingo; Adam, Elisabeth; Adrião, Diana; Ageel, Saleh Al; Ain, Quratul; Ainscough, Kate; Ait Hssain, Ali; Ait Tamlihat, Younes; Akimoto, Takako; Akmal, Ernita; Al Qasim, Eman; Alalqam, Razi; Alam, Tanvir; Al-dabbous, Tala; Alegre, Cynthia; Alessi, Marta; Alex, Beatrice; Alexandre, Kévin; Al-Fares, Abdulrahman; Alfoudri, Huda; Ali Shah, Naseem; Alidjnou, Kazali Enagnon; Aliudin, Jeffrey; Allavena, Clotilde; Allou, Nathalie; Altaf, Aneela; Alves, João Melo; Alves, João; Alves, Rita; Amaral, Maria; Amira, Nur; Ammerlaan, Heidi; Amorim Beltrão, Beatriz Amorim; Ampaw, Phoebe; Andini, Roberto; Andrejak, Claire; Angheben, Andrea; Angoulvant, François; Ansart, Séverine; Anthonidass, Sivanesen; Antonelli, Massimo; Antunes de Brito, Carlos Alexandre; Anwar, Kazi Rubayet; Apriyana, Ardiyan; Arabi, Yaseen; Aragao, Irene; Arancibia, Francisco; Araújo-Mariz, Carolline; Arcadipane, Antonio; Archambault, Patrick; Arenz, Lukas; Arlet, Jean-Benoît; Arnold-Day, Christel; Aroca, Ana; Arora, Lovkesh; Arora, Rakesh; Artaud-Macari, Elise; Aryal, Diptesh; Asaki, Motohiro; Asensio, Angel; Ashraf, Muhammad; Asim, Mohammad; Assie, Jean Baptiste; Asyraf, Amirul; Atique, Anika; Attanyake, AM Udara Lakshan; Auchabie, Johann; Aumaitre, Hugues; Auvet, Adrien; Azemar, Laurène; Azoulay, Cecile; Bach, Benjamin; Bachelet, Delphine; Badr, Claudine; Baig, Nadia; Baillie, J. Kenneth; Bak, Erica; Bakakos, Agamemnon; Bakar, Nazreen Abu; Bal, Andriy; Balakrishnan, Mohanaprasanth; Bani-Sadr, Firouzé; Barbalho, Renata; Barbosa, Nicholas Yuri; Barclay, Wendy S.; Barnett, Saef Umar; Barnikel, Michaela; Barrasa, Helena; Barrelet, Audrey; Barrigoto, Cleide; Bartoli, Marie; Bartone, Cheryl; Baruch, Joaquín; Basmaci, Romain; Basri, Muhammad Fadhli Hassin; Battaglini, Denise; Bauer, Jules; Bautista Rincon, Diego Fernando; Bazan Dow, Denisse; Beane, Abigail; Bedossa, Alexandra; Bee, Ker Hong; Behilill, Sylvie; Beishuizen, Albertus; Beljantsev, Aleksandr; Bellemare, David; Beltrame, Anna; Beluze, Marine; Benech, Nicolas; Benjiman, Lionel Eric; Benkerrou, Dehbia; Bennett, Suzanne; Bento, Luís; Berdal, Jan-Erik; Bergeaud, Delphine; Bernal Sobrino, José Luis; Bertoli, Giulia; Bertolino, Lorenzo; Bessis, Simon; Betz, Adam; Bevilcaqua, Sybille; Bezulier, Karine; Bhatt, Amar; Bhavsar, Krishna; Bianchi, Isabella; Bianco, Claudia; Bidin, Farah Nadiah; Bikram Singh, Moirangthem; Bin Humaid, Felwa; Bin Kamarudin, Mohd Nazlin; Bissuel, François; Biston, Patrick; Bitker, Laurent; Blanco-Schweizer, Pablo; Blier, Catherine; Bloos, Frank; Blot, Mathieu; Blumberg, Lucille; Boccia, Filomena; Bodenes, Laetitia; Bogaarts, Alice; Bogaert, Debby; Boivin, Anne-Hélène; Bolze, Pierre-Adrien; Bompart, François; Borges, Diogo; Borie, Raphaël; Bosse, Hans Martin; Botelho-Nevers, Elisabeth; Bouadma, Lila; Bouchaud, Olivier; Bouchez, Sabelline; Bouhmani, Dounia; Bouhour, Damien; Bouiller, Kévin; Bouillet, Laurence; Bouisse, Camile; Boureau, Anne-Sophie; Bouscambert, Maude; Bousquet, Aurore; Bouziotis, Jason; Boxma, Bianca; Boyer-Besseyre, Marielle; Boylan, Maria; Bozza, Fernando Augusto; Brack, Matthew; Braconnier, Axelle; Braga, Cynthia; Brandenburger, Timo; Brás Monteiro, Filipa; Brazzi, Luca; Breen, Dorothy; Breen, Patrick; Brickell, Kathy; Broadley, Tessa; Browne, Alex; Brozzi, Nicolas; Brusse-Keizer, Marjolein; Buchtele, Nina; Buesaquillo, Christian; Bugaeva, Polina; Buisson, Marielle; Burhan, Erlina; Burrell, Aidan; Bustos, Ingrid G.; Butnaru, Denis; Cabie, André; Cabral, Susana; Caceres, Eder; Cadoz, Cyril; Callahan, Mia; Calligy, Kate; Calvache, Jose Andres; Camões, João; Campana, Valentine; Campbell, Paul; Canepa, Cecilia; Cantero, Mireia; Caraux-Paz, Pauline; Cárcel, Sheila; Cardellino, Chiara Simona; Cardoso, Filipa; Cardoso, Filipe; Cardoso, Nelson; Cardoso, Sofia; Carelli, Simone; Carlier, Nicolas; Carmoi, Thierry; Carney, Gayle; Carpenter, Chloe; Carqueja, Inês; Carret, Marie-Christine; Carrier, François Martin; Carson, Gail; Casanova, Maire-Laure; Cascão, Mariana; Casimiro, José; Cassandra, Bailey; Castañeda, Silvia; Castanheira, Nidyanara; Castor-Alexandre, Guylaine; Castrillón, Henry; Castro, Ivo; Catarino, Ana; Catherine, François-Xavier; Cattaneo, Paolo; Cavalin, Roberta; Cavalli, Giulio Giovanni; Cavayas, Alexandros; Ceccato, Adrian; Cervantes-Gonzalez, Minerva; Chair, Anissa; Chakveatze, Catherine; Chan, Adrienne; Chand, Meera; Chantalat Auger, Christelle; Chapplain, Jean-Marc; Chas, Julie; Chaudary, Mobin; Chávez Iñiguez, Jonathan Samuel; Chen, Anjellica; Chen, Yih-Sharng; Cheng, Matthew Pellan; Cheret, Antoine; Chiarabini, Thibault; Chica, Julian; Chidambaram, Suresh Kumar; Chin-Tho, Leong; Chirouze, Catherine; Chiumello, Davide; Cho, Hwa Jin; Cho, Sung-Min; Cholley, Bernard; Chopin, Marie-Charlotte; Chow, Yock Ping; Chow, Ting Soo; Chua, Hiu Jian; Chua, Jonathan; Cidade, Jose Pedro; Cisneros Herreros, Jose Miguel; Citarella, Barbara Wanjiru; Ciullo, Anna; Clarke, Jennifer; Claure Del Granado, Rolando; Clohisey, Sara; Coca, Necsoi; Codan, Cassidy; Cody, Caitriona; Coelho, Alexandra; Colin, Gwenhaël; Collins, Michael; Colombo, Sebastiano Maria; Combs, Pamela; Conrad, Anne; Contreras, Sofía; Conway, Elaine; Cooke, Graham S.; Copland, Mary; Cordel, Hugues; Corley, Amanda; Cormican, Sarah; Cornelis, Sabine; Cornet, Alexander Daniel; Corpuz, Arianne Joy; Cortegiani, Andrea; Corvaisier, Grégory; Couffignal, Camille; Couffin-Cadiergues, Sandrine; Courtois, Roxane; Cousse, Stéphanie; Crepy D’Orleans, Charles; Croonen, Sabine; Crowl, Gloria; Crump, Jonathan; Cruz, Claudina; Cruz Bermúdez, Juan Luis; Cruz Rojo, Jaime; Csete, Marc; Cucino, Alberto; Cullen, Caroline; Cummings, Matthew; Curley, Gerard; Curlier, Elodie; Custodio, Paula; da Silva Filipe, Ana; Da Silveira, Charlene; Dabaliz, Al-Awwab; Dagens, Andrew; Dalton, Heidi; D’Amico, Federico; Daneman, Nick; Daniel, Corinne; Dankwa, Emmanuelle A; Dantas, Jorge; D’Aragon, Frédérick; de Boer, Mark; de Mendoza, Diego; De Montmollin, Etienne; de Oliveira França, Rafael Freitas; de Pinho Oliveira, Ana Isabel; De Rosa, Rosanna; de Silva, Thushan; de Vries, Peter; Deacon, Jillian; Dean, David; Debard, Alexa; Debray, Marie-Pierre; DeCastro, Nathalie; Dechert, William; Deconninck, Lauren; Decours, Romain; Defous, Eve; Delacroix, Isabelle; Delaveuve, Eric; Delavigne, Karen; Delfos, Nathalie M.; Deligiannis, Ionna; Dell’Amore, Andrea; Delmas, Christelle; Delobel, Pierre; Delsing, Corine; Demonchy, Elisa; Denis, Emmanuelle; Deplanque, Dominique; Depuydt, Pieter; Desai, Mehul; Descamps, Diane; Desvallée, Mathilde; Dewayanti, Santi; Diallo, Alpha; Diamantis, Sylvain; Dias, André; Diaz, Rodrigo; Diaz, Juan Jose; Diaz, Priscila; Didier, Kévin; Diehl, Jean-Luc; Dieperink, Wim; Dimet, Jérôme; Dinot, Vincent; Diop, Fara; Diouf, Alphonsine; Dishon, Yael; Djossou, Félix; Docherty, Annemarie B.; Dondorp, Arjen M; Dong, Andy; Donnelly, Christl A.; Donnelly, Maria; Donohue, Chloe; Doran, Peter; Dorival, Céline; D’Ortenzio, Eric; Douglas, James Joshua; Douma, Renee; Dournon, Nathalie; Downer, Triona; Downing, Mark; Drake, Tom; Driscoll, Aoife; Dryden, Murray; Duarte Fonseca, Claudio; Dubee, Vincent; Dubos, François; Ducancelle, Alexandre; Duculan, Toni; Dudman, Susanne; Duggal, Abhijit; Dunand, Paul; Dunning, Jake; Duplaix, Mathilde; Durante-Mangoni, Emanuele; Durham III, Lucian; Dussol, Bertrand; Duthoit, Juliette; Duval, Xavier; Dyrhol-Riise, Anne Margarita; Ean, Sim Choon; Echeverria-Villalobos, Marco; Egan, Siobhan; Eira, Carla; El Sanharawi, Mohammed; Elapavaluru, Subbarao; Elharrar, Brigitte; Ellerbroek, Jacobien; Ellis, Rachael; Eloy, Philippine; Elshazly, Tarek; Enderle, Isabelle; Endo, Tomoyuki; Eng, Chan Chee; Engelmann, Ilka; Enouf, Vincent; Epaulard, Olivier; Escher, Martina; Esperatti, Mariano; Esperou, Hélène; Esposito-Farese, Marina; Estevão, João; Etienne, Manuel; Ettalhaoui, Nadia; Everding, Anna Greti; Evers, Mirjam; Fabre, Isabelle; Fabre, Marc; Faheem, Amna; Fahy, Arabella; Fairfield, Cameron J.; Faria, Pedro; Farooq, Ahmed; Farshait, Nataly; Fateena, Hanan; Fatoni, Arie Zainul; Faure, Karine; Favory, Raphaël; Fayed, Mohamed; Feely, Niamh; Fernandes, Jorge; Fernandes, Marília; Fernandes, Susana; Ferrand, François-Xavier; Ferrand Devouge, Eglantine; Ferrão, Joana; Ferraz, Mário; Ferreira, Benigno; Ferreira, Sílvia; Ferrer-Roca, Ricard; Ferriere, Nicolas; Ficko, Céline; Figueiredo-Mello, Claudia; Fiorda, Juan; Flament, Thomas; Flateau, Clara; Fletcher, Tom; Florio, Letizia Lucia; Flynn, Brigid; Foley, Claire; Fomin, Victor; Fonseca, Tatiana; Fontela, Patricia; Forsyth, Simon; Foster, Denise; Foti, Giuseppe; Fourn, Erwan; Fowler, Robert A.; Fraher, Dr Marianne; Franch-Llasat, Diego; Fraser, Christophe; Fraser, John; Freire, Marcela Vieira; Freitas Ribeiro, Ana; Friedrich, Caren; Fritz, Ricardo; Fry, Stéphanie; Fuentes, Nora; Fukuda, Masahiro; Gaborieau, Valérie; Gaci, Rostane; Gagliardi, Massimo; Gagnard, Jean-Charles; Gagné, Nathalie; Gagneux-Brunon, Amandine; Gaião, Sérgio; Gail Skeie, Linda; Gallagher, Phil; Gallego Curto, Elena; Gamble, Carrol; Gani, Yasmin; Garan, Arthur; Garcia, Rebekha; García Barrio, Noelia; Garcia-Gallo, Esteban; Garimella, Navya; Garot, Denis; Garrait, Valérie; Gauli, Basanta; Gault, Nathalie; Gavin, Aisling; Gavrylov, Anatoliy; Gaymard, Alexandre; Gebauer, Johannes; Geraud, Eva; Gerbaud Morlaes, Louis; Germano, Nuno; Ghosn, Jade; Giani, Marco; Giaquinto, Carlo; Gibson, Jess; Gigante, Tristan; Gilg, Morgane; Giordano, Guillermo; Girvan, Michelle; Gissot, Valérie; Gitahi, Judy; Giwangkancana, Gezy; Glikman, Daniel; Glybochko, Petr; Gnall, Eric; Goco, Geraldine; Goehringer, François; Goepel, Siri; Goffard, Jean-Christophe; Goh, Jin Yi; Golob, Jonathan; Gomes, Rui; Gomez, Kyle; Gómez-Junyent, Joan; Gominet, Marie; Gonzalez, Alicia; Gorenne, Isabelle; Goubert, Laure; Goujard, Cécile; Goulenok, Tiphaine; Grable, Margarite; Graf, Jeronimo; Grandin, Edward Wilson; Granier, Pascal; Grasselli, Giacomo; Grazioli, Lorenzo; Green, Christopher A.; Greenhalf, William; Greffe, Segolène; Grieco, Domenico Luca; Griffee, Matthew; Griffiths, Fiona; Grigoras, Ioana; Groenendijk, Albert; Grosse Lordemann, Anja; Gruner, Heidi; Gu, Yusing; Guarracino, Fabio; Guedj, Jérémie; Guego, Martin; Guellec, Dewi; Guerguerian, Anne-Marie; Guerreiro, Daniela; Guery, Romain; Guillaumot, Anne; Guilleminault, Laurent; Guimarães de Castro, Maisa; Guimard, Thomas; Haalboom, Marieke; Haber, Daniel; Hachemi, Ali; Hadri, Nadir; Haidash, Olena; Haider, Saeeda; Haidri, Fakhir; Hakak, Sheeba; Hall, Adam; Hall, Matthew; Halpin, Sophie; Hamer, Ansley; Hamidfar, Rebecca; Hammond, Terese; Han, Lim Yuen; Haniffa, Rashan; Hao, Kok Wei; Hardwick, Hayley; Harrison, Ewen M.; Harrison, Janet; Harrison, Samuel Bernard Ekow; Hartman, Alan; Hashmi, Madiha; Hayat, Muhammad; Hays, Leanne; Heerman, Jan; Heggelund, Lars; Hendry, Ross; Hennessy, Martina; Henriquez-Trujillo, Aquiles; Hentzien, Maxime; Herekar, Fivzia; Hernandez-Montfort, Jaime; Herr, Daniel; Hershey, Andrew; Hesstvedt, Liv; Hidayah, Astarini; Higgins, Eibhilin; Higgins, Dawn; Hinton, Samuel; Hiraiwa, Hiroaki; Hirkani, Haider; Hitoto, Hikombo; Ho, Antonia Ying Wai; Ho, Yi Bin; Hoctin, Alexandre; Hoffmann, Isabelle; Hoh, Wei Han; Hoiting, Oscar; Holt, Rebecca; Holter, Jan Cato; Horby, Peter; Horcajada, Juan Pablo; Hoshino, Koji; Hoshino, Kota; Houas, Ikram; Hough, Catherine L.; Houltham, Stuart; Hsu, Jimmy Ming-Yang; Hulot, Jean-Sébastien; Hussain, Iqbal; Ijaz, Samreen; Illes, Hajnal-Gabriela; Imbert, Patrick; Imran, Mohammad; Imran Sikander, Rana; Inácio, Hugo; Infante Dominguez, Carmen; Ing, Yun Sii; Iosifidis, Elias; Ippolito, Mariachiara; Isgett, Sarah; Ishani, Palliya Guruge Pramodya Ishani; Isidoro, Tiago; Ismail, Nadiah; Isnard, Margaux; Itai, Junji; Ivulich, Daniel; Jaafar, Danielle; Jaafoura, Salma; Jabot, Julien; Jackson, Clare; Jamieson, Nina; Jaquet, Pierre; Jassat, Waasila; Jaud-Fischer, Coline; Jaureguiberry, Stéphane; Javidfar, Jeffrey; Jawad, Issrah; Jaworsky, Denise; Jayakumar, Devachandran; Jego, Florence; Jelani, Anilawati Mat; Jenum, Synne; Jimbo-Sotomayor, Ruth; Job, van der Palen; Joe, Ong Yiaw; Jorge García, Ruth N.; Joseph, Cédric; Joseph, Mark; Joshi, Swosti; Jourdain, Mercé; Jouvet, Philippe; June, Jennifer; Jung, Anna; Jung, Hanna; Juzar, Dafsah; Kafif, Ouifiya; Kaguelidou, Florentia; Kaisbain, Neerusha; Kaleesvran, Thavamany; Kali, Sabina; Kalicinska, Alina; Kalomoiri, Smaragdi; Kamaluddin, Muhammad Aisar Ayadi; Kamaruddin, Zul Amali Che; Kamarudin, Nadiah; Kandamby, Darshana Hewa; Kandel, Chris; Kang, Kong Yeow; Kant, Ravi; Kanwal, Darakhshan; Kanyawati, Dyah; Karpayah, Pratap; Karsies, Todd; Kartsonaki, Christiana; Kasugai, Daisuke; Kataria, Anant; Katz, Kevin; Kaur Johal, Simreen; Kawasaki, Tatsuya; Kay, Christy; Keating, Sean; Kedia, Pulak; Kelly, Andrea; Kelly, Sadie; Kennedy, Ryan; Kennon, Kalynn; Kerroumi, Younes; Kestelyn, Evelyne; Khalid, Imrana; Khalid, Osama; Khalil, Antoine; Khan, Coralie; Khan, Irfan; Khanal, Sushil; Kherajani, Krish; Kho, Michelle E; Khoo, Denisa; Khoo, Ryan; Khoo, Saye; Khoso, Nasir; Kiat, Khor How; Kida, Yuri; Kiiza, Peter; Kildal, Anders Benjamin; Kim, Jae Burm; Kimmoun, Antoine; Kindgen-Milles, Detlef; King, Alexander H.; Kitamura, Nobuya; Klenerman, Paul; Klont, Rob; Kloumann Bekken, Gry; Knight, Stephen; Kobbe, Robin; Kodippily, Chamira; Kohns Vasconcelos, Malte; Koirala, Sabin; Komatsu, Mamoru; Korten, Volkan; Kosgei, Caroline; Kpangon, Arsène; Krawczyk, Karolina; Krishnan, Sudhir; Krishnan, Vinothini; Kruglova, Oksana; Kumar, Deepali; Kumar, Ganesh; Kumar, Ashok; Kumar, Mukesh; Kumar Tirupakuzhi Vijayaraghavan, Bharath; Kumar Vecham, Pavan; Kurtzman, Ethan; Kusumastuti, Neurinda Permata; Kutsogiannis, Demetrios; Kutsyna, Galyna; Kyriakoulis, Konstantinos; Lachatre, Marie; Lacoste, Marie; Laffey, John G.; Lagrange, Marie; Laine, Fabrice; Lairez, Olivier; Lalueza Blanco, Antonio; Lambert, Marc; Lamontagne, François; Langelot-Richard, Marie; Langlois, Vincent; Lantang, Eka Yudha; Lanza, Marina; Laouénan, Cédric; Laribi, Samira; Lariviere, Delphine; Lasry, Stéphane; Lath, Sakshi; Latif, Naveed; Launay, Odile; Laureillard, Didier; Lavie-Badie, Yoan; Law, Andrew; Lawrence, Cassie; Le, Minh; Le Bihan, Clément; Le Bris, Cyril; Le Falher, Georges; Le Fevre, Lucie; Le Hingrat, Quentin; Le Maréchal, Marion; Le Mestre, Soizic; Le Moal, Gwenaël; Le Moing, Vincent; Le Nagard, Hervé; Le Turnier, Paul; Leal, Ema; Leal Santos, Marta; Lee, Biing Horng; Lee, Heng Gee; Lee, Su Hwan; Lee, James; Lee, Todd C.; Lee, Yi Lin; Leeming, Gary; Lefebvre, Bénédicte; Lefebvre, Laurent; Lefevre, Benjamin; LeGac, Sylvie; Lelievre, Jean-Daniel; Lellouche, François; Lemaignen, Adrien; Lemee, Véronique; Lemeur, Anthony; Lemmink, Gretchen; Lene, Ha Sha; León, Rafael; Leone, Marc; Leone, Michela; Lepiller, Quentin; Lescure, François-Xavier; Lesens, Olivier; Lesouhaitier, Mathieu; Levy, Bruno; Levy, Yves; Levy-Marchal, Claire; Lewandowska, Katarzyna; L’Her, Erwan; Li Bassi, Gianluigi; Liang, Janet; Liaquat, Ali; Liegeon, Geoffrey; Lim, Kah Chuan; Lim, Wei Shen; Lima, Chantre; Lina, Bruno; Lina, Lim; Lind, Andreas; Lingas, Guillaume; Lion-Daolio, Sylvie; Lissauer, Samantha; Liu, Keibun; Livrozet, Marine; Lizotte, Patricia; Loforte, Antonio; Lolong, Navy; Loon, Leong Chee; Lopes, Diogo; Lopez-Colon, Dalia; Loschner, Anthony L.; Loubet, Paul; Loufti, Bouchra; Louis, Guillame; Lourenco, Silvia; Lovelace-Macon, Lara; Low, Lee Lee; Lowik, Marije; Loy, Jia Shyi; Lucet, Jean Christophe; Lumbreras Bermejo, Carlos; Luna, Carlos M.; Lungu, Olguta; Luong, Liem; Luque, Nestor; Luton, Dominique; Lwin, Nilar; Lyons, Ruth; Maasikas, Olavi; Mabiala, Oryane; MacDonald, Sarah; MacDonald, Samual; Machado, Moïse; Macheda, Gabriel; Macias Sanchez, Juan; Madhok, Jai; Maestro de la Calle, Guillermo; Mahieu, Rafael; Mahy, Sophie; Maia, Ana Raquel; Maier, Lars Siegfrid; Maillet, Mylène; Maitre, Thomas; Malfertheiner, Maximilian; Malik, Nadia; Maltez, Fernando; Malvy, Denis; Manda, Victoria; Mandei, Jose M.; Mandelbrot, Laurent; Mangal, Kishore; Mankikian, Julie; Manning, Edmund; Manuel, Aldric; Maria Sant’Ana Malaque, Ceila; Marino, Daniel; Marino, Flávio; Markowicz, Samuel; Maroun Eid, Charbel; Marques, Ana; Marquis, Catherine; Marsh, Brian; Marshall, John; Martelli, Celina Turchi; Martin, Dori-Ann; Martin, Emily; Martin-Blondel, Guillaume; Martinelli, Alessandra; Martin-Loeches, Ignacio; Martinot, Martin; Martin-Quiros, Alejandro; Martins, Ana; Martins, João; Martins, Nuno; Martins Rego, Caroline; Martucci, Gennaro; Martynenko, Olga; Marwali, Eva Miranda; Marzukie, Marsilla; Masa Jimenez, Juan Fernado; Maslove, David; Mason, Phillip; Mason, Sabina; Masood, Sobia; Masood, Sobia; Mat Nor, Basri; Matan, Moshe; Mathew, Meghena; Mathieu, Daniel; Mattei, Mathieu; Matulevics, Romans; Maulin, Laurence; Maynar, Javier; Mazzoni, Thierry; Mc Evoy, Natalie; McArthur, Colin; McCarthy, Aine; McCarthy, Anne; McCloskey, Colin; McConnochie, Rachael; McDermott, Sherry; McDonald, Sarah; McElwee, Samuel; McGeer, Allison; McKay, Chris; McKeown, Johnny; McLean, Kenneth A.; McNicholas, Bairbre; Meaney, Edel; Mear-Passard, Cécile; Mechlin, Maggie; Meher, Maqsood; Mehkri, Omar; Mele, Ferruccio; Melo, Luis; Memon, Kashif; Mendes, Joao Joao; Menkiti, Ogechukwu; Menon, Kusum; Mentré, France; Mentzer, Alexander J.; Mercier, Emmanuelle; Mercier, Noémie; Merckx, Antoine; Mergeay-Fabre, Mayka; Mergler, Blake; Merson, Laura; Mesquita, António; Meybeck, Agnès; Meyer, Dan; Meynert, Alison M.; Meysonnier, Vanina; Meziane, Amina; Mezidi, Mehdi; Michelagnoli, Giuliano; Michelanglei, Céline; Michelet, Isabelle; Mihelis, Efstathia; Mihnovit, Vladislav; Miranda-Maldonado, Hugo; Misnan, Nor Arisah; Mohamed, Nik Nur Eliza; Mohamed, Tahira Jamal; Moin, Asma; Molina, David; Molinos, Elena; Mone, Mary; Monteiro, Agostinho; Montes, Claudia; Montrucchio, Giorgia; Moore, Sarah; Moore, Shona C.; Morales Cely, Lina; Moro, Lucia; Morocho Tutillo, Diego Rolando; Morton, Ben; Motherway, Catherine; Motos, Ana; Mouquet, Hugo; Mouton Perrot, Clara; Moyet, Julien; Mudara, Caroline; Mufti, Aisha Kalsoom; Muh, Ng Yong; Muhamad, Dzawani; Mullaert, Jimmy; Muller, Fredrik; Müller, Karl Erik; Munblit, Daniel; Muneeb, Syed; Munir, Nadeem; Murris, Marlène; Murthy, Srinivas; Muyandy, Gugapriyaa; Myrodia, Dimitra Melia; N, Namra; Nagpal, Dave; Nagrebetsky, Alex; Nasim Khan, Rashid; Neant, Nadège; Neb, Holger; Nekliudov, Nikita A; Neto, Raul; Neumann, Emily; Neves, Bernardo; Ng, Wing Yiu; Ng, Pauline Yeung; Nghi, Anthony; Nguyen, Duc; Ni Choileain, Orna; Nichol, Alistair; Nitayavardhana, Prompak; Nonas, Stephanie; Noordin, Nurul Amani Mohd; Noret, Marion; Norharizam, Nurul Faten Izzati; Norman, Lisa; Notari, Alessandra; Noursadeghi, Mahdad; Nowicka, Karolina; Nowinski, Adam; Nseir, Saad; Nunez, Jose I; Nurnaningsih, Nurnaningsih; Nyamankolly, Elsa; Occhipinti, Giovanna; O’Donnell, Max; Ogston, Tawnya; Ogura, Takayuki; Oh, Tak-Hyuk; O’Hearn, Katie; Ohshimo, Shinichiro; Ołdakowska, Agnieszka; Oliveira, João; Oliveira, Larissa; Olliaro, Piero L.; O’Neil, Conar; Ong, David S.Y.; Ong, Jee Yan; Oosthuyzen, Wilna; Opavsky, Anne; Openshaw, Peter; Orakzai, Saijad; Orozco-Chamorro, Claudia Milena; Orquera, Andrés; Ortoleva, Jamel; Osatnik, Javier; Othman, Siti Zubaidah; Ouamara, Nadia; Ouissa, Rachida; Owyang, Clark; Oziol, Eric; Pabasara, H M Upulee; Pagadoy, Maïder; Pages, Justine; Palacios, Amanda; Palacios, Mario; Palmarini, Massimo; Panarello, Giovanna; Panda, Prasan Kumar; Paneru, Hem; Pang, Lai Hui; Panigada, Mauro; Pansu, Nathalie; Papadopoulos, Aurélie; Parke, Rachael; Parker, Melissa; Parra, Briseida; Parrini, Vieri; Pasha, Taha; Pasquier, Jérémie; Pastene, Bruno; Patauner, Fabian; Patel, Drashti; Patel, Junaid; Pathmanathan, Mohan Dass; Patrão, Luís; Patricio, Patricia; Patrier, Juliette; Patterson, Lisa; Pattnaik, Rajyabardhan; Paul, Christelle; Paul, Mical; Paulos, Jorge; Paxton, William A.; Payen, Jean-François; Peariasamy, Kalaiarasu; Pedrera Jiménez, Miguel; Peek, Giles J.; Peelman, Florent; Peiffer-Smadja, Nathan; Peigne, Vincent; Pejkovska, Mare; Pelosi, Paolo; Peltan, Ithan D.; Pereira, Rui; Perez, Daniel; Periel, Luis; Perpoint, Thomas; Pesenti, Antonio; Pestre, Vincent; Petrou, Lenka; Petrov-Sanchez, Ventzislava; Pettersen, Frank Olav; Peytavin, Gilles; Pharand, Scott; Piagnerelli, Michael; Picard, Walter; Picone, Olivier; Piero, Maria de; Pierobon, Carola; Piersma, Djura; Pimentel, Carlos; Pinto, Raquel; Pires, Catarina; Pironneau, Isabelle; Piroth, Lionel; Pius, Riinu; Piva, Simone; Plantier, Laurent; Plotkin, Daniel; Png, Hon Shen; Poissy, Julien; Pokeerbux, Ryadh; Pokorska-Śpiewak, Maria; Poli, Sergio; Pollakis, Georgios; Ponscarme, Diane; Popielska, Jolanta; Porto, Diego B. ; Post, Andra-Maris; Postma, Douwe F.; Povoa, Pedro; Póvoas, Diana; Powis, Jeff; Prapa, Sofia; Preau, Sébastien; Prebensen, Christian; Preiser, Jean-Charles; Prinssen, Anton; Pritchard, Mark G.; Priyadarshani, Gamage Dona Dilanthi; Proença, Lucia; Puéchal, Oriane; Pujo Semedi, Bambang; Pulicken, Mathew; Purcell, Gregory; Quesada, Luisa; Quinones-Cardona, Vilmaris; Quirós González, Víctor; Quist-Paulsen, Else; Quraishi, Mohammed; Rabaud, Christian; Rabindrarajan, Ebenezer; Rafael, Aldo; Rafiq, Marie; Ragazzo, Gabrielle; Rahman, Ahmad Kashfi Haji Ab; Rahman, Rozanah Abd; Rahutullah, Arsalan; Rainieri, Fernando; Rajahram, Giri Shan; Ralib, Azrina; Ramakrishnan, Nagarajan; Ramanathan, Kollengode; Ramli, Ahmad Afiq; Rammaert, Blandine; Ramos, Grazielle Viana; Rana, Asim; Rapp, Christophe; Rashan, Aasiyah; Rashan, Thalha; Rasheed, Ghulam; Rasmin, Menaldi; Rätsep, Indrek; Rau, Cornelius; Ravi, Tharmini; Raza, Ali; Real, Andre; Rebaudet, Stanislas; Redl, Sarah; Reeve, Brenda; Rehan, Ali; Rehman, Attaur; Reid, Liadain; Reikvam, Dag Henrik; Reis, Renato; Rello, Jordi; Remppis, Jonathan; Remy, Martine; Ren, Hongru; Renk, Hanna; Resende, Liliana; Resseguier, Anne-Sophie; Revest, Matthieu; Rewa, Oleksa; Reyes, Luis Felipe; Reyes, Tiago; Ribeiro, Maria Ines; Richardson, David; Richardson, Denise; Richier, Laurent; Ridzuan, Siti Nurul Atikah Ahmad; Riera, Jordi; Rios, Ana L; Rishu, Asgar; Rispal, Patrick; Risso, Karine; Rivera Nuñez, Maria Angelica; Rizer, Nicholas; Robba, Chiara; Roberto, André; Robertson, David L.; Robineau, Olivier; Roche-Campo, Ferran; Rodari, Paola; Rodeia, Simão; Rodriguez Abreu, Julia; Roessler, Bernhard; Roger, Claire; Roger, Pierre-Marie; Roilides, Emmanuel; Rojek, Amanda; Romaru, Juliette; Roncon-Albuquerque Jr, Roberto; Roriz, Mélanie; Rosa-Calatrava, Manuel; Rose, Michael; Rosenberger, Dorothea; Roslan, Nurul Hidayah Mohammad; Rossanese, Andrea; Rossetti, Matteo; Rossignol, Bénédicte; Rossignol, Patrick; Rousset, Stella; Roy, Carine; Roze, Benoît; Rusmawatiningtyas, Desy; Russell, Clark D.; Ryckaert, Steffi; Rygh Holten, Aleksander; Saba, Isabela; Sadaf, Sairah; Sadat, Musharaf; Sahraei, Valla; Saint-Gilles, Maximilien; Sakiyalak, Pranya; Salahuddin, Nawal; Salazar, Leonardo; Saleem, Jodat; Sales, Gabriele; Sallaberry, Stéphane; Salmon Gandonniere, Charlotte; Salvator, Hélène; Sanchez, Olivier; Sánchez Choez, Xavier; Sanchez-Miralles, Angel; Sancho-Shimizu, Vanessa; Sandhu, Gyan; Sandrine, Pierre-François; Sandulescu, Oana; Santos, Marlene; Sarfo-Mensah, Shirley; Sarmento Banheiro, Bruno; Sarmiento, Iam Claire E.; Sarton, Benjamine; Satya, Ankana; Satyapriya, Sree; Satyawati, Rumaisah; Savvidou, Parthena; Saw, Yen Tsen; Schaffer, Justin; Schermer, Tjard; Scherpereel, Arnaud; Schneider, Marion; Schroll, Stephan; Schwameis, Michael; Scott, Janet T.; Scott-Brown, James; Sedillot, Nicholas; Seitz, Tamara; Selvanayagam, Jaganathan; Selvarajoo, Mageswari; Semaille, Caroline; Semple, Malcolm G.; Senian, Rasidah Bt; Senneville, Eric; Sepulveda, Claudia; Sequeira, Filipa; Sequeira, Tânia; Serpa Neto, Ary; Serrano Balazote, Pablo; Shadowitz, Ellen; Shahidan, Syamin Asyraf; Shamsah, Mohammad; Sharma, Pratima; Shaw, Catherine A.; Shaw, Victoria; Sheharyar, Ashraf; Shetty, Rohan; Shi, Haixia; Shiban, Nisreen; Shiekh, Mohiuddin; Shiga, Takuya; Shime, Nobuaki; Shimizu, Hiroaki; Shimizu, Keiki; Shimizu, Naoki; Shrapnel, Sally; Shum, Hoi Ping; Si Mohammed, Nassima; Siang, Ng Yong; Sibiude, Jeanne; Siddiqui, Atif; Sigfrid, Louise; Sillaots, Piret; Silva, Catarina; Silva, Maria Joao; Silva, Rogério; Sim Lim Heng, Benedict; Sin, Wai Ching; Singh, Budha Charan ; Sitompul, Pompini Agustina; Sivam, Karisha; Skogen, Vegard; Smith, Sue; Smood, Benjamin; Smyth, Michelle; Snacken, Morgane; So, Dominic; Soh, Tze Vee; Solis, Monserrat; Solomon, Joshua; Solomon, Tom; Somers, Emily; Sommet, Agnès; Song, Myung Jin; Song, Rima; Song, Tae; Sonntagbauer, Michael; Soom, Azlan Mat; Sotto, Alberto; Soum, Edouard; Sousa, Ana Chora; Sousa, Marta; Sousa Uva, Maria; Souza-Dantas, Vicente; Sperry, Alexandra; Spinuzza, Elisabetta; Sri Darshana, B. P. Sanka Ruwan; Sriskandan, Shiranee; Stabler, Sarah; Staudinger, Thomas; Stecher, Stephanie-Susanne; Steinsvik, Trude; Stienstra, Ymkje; Stiksrud, Birgitte; Streinu-Cercel, Anca; Streinu-Cercel, Adrian; Strudwick, Samantha; Stuart, Ami; Stuart, David; Suen, Gabriel; Suen, Jacky Y.; Sultana, Asfia; Summers, Charlotte; Suppiah, Deepashankari; Surovcová, Magdalena; Svistunov, Andrey A; Syahrin, Sarah; Syrigos, Konstantinos; Sztajnbok, Jaques; Szuldrzynski, Konstanty; Tabrizi, Shirin; Taccone, Fabio S.; Tagherset, Lysa; Taib, Shahdattul Mawarni; Talarek, Ewa; Taleb, Sara; Talsma, Jelmer; Tampubolon, Maria Lawrensia; Tan, Le Van; Tan, Kim Keat; Tan, Yan Chyi; Tanaka, Clarice; Tanaka, Hiroyuki; Tanaka, Taku; Taniguchi, Hayato; Tanveer, Hussain; Taqdees, Huda; Taqi, Arshad; Tardivon, Coralie; Tattevin, Pierre; Taufik, M Azhari; Tedder, Richard S.; Tee, Tze Yuan; Teixeira, João; Tejada, Sofia; Tellier, Marie-Capucine; Teoh, Sze Kye; Teotonio, Vanessa; Téoulé, François; Terpstra, Pleun; Terrier, Olivier; Terzi, Nicolas; Tessier-Grenier, Hubert; Tey, Adrian; Thabit, Alif Adlan Mohd; Tham, Zhang Duan; Thangavelu, Suvintheran; Thibault, Vincent; Thiberville, Simon-Djamel; Thill, Benoît; Thirumanickam, Jananee; Thompson, Shaun; Thomson, Emma C.; Thomson, David; Thurai, Surain Raaj Thanga; Thuy, Duong Bich; Thwaites, Ryan S.; Tieroshyn, Vadim; Timashev, Peter S; Timsit, Jean-François; Tissot, Noémie; Toh, Jordan Zhien Yang; Toki, Maria; Tolppa, Timo; Tonby, Kristian; Tonnii, Sia Loong; Torres, Antoni; Torres, Margarida; Torres Santos-Olmo, Rosario Maria; Torres-Zevallos, Hernando; Trapani, Tony; Treoux, Théo; Trieu, Huynh Trung; Tripathy, Swagata; Tromeur, Cécile; Trontzas, Ioannis; Trouillon, Tiffany; Truong, Jeanne; Tual, Christelle; Tubiana, Sarah; Tuite, Helen; Turmel, Jean-Marie; Turtle, Lance C.W.; Tveita, Anders; Twardowski, Pawel; Uchiyama, Makoto; Udayanga, PG Ishara; Udy, Andrew; Ullrich, Roman; Umer, Zubair; Uribe, Alberto; Usman, Asad; Val-Flores, Luís; Valle, Ana Luiza; Valran, Amélie; Van de Velde, Stijn; van den Berge, Marcel; Van der Feltz, Machteld; van der Valk, Paul; Van Der Vekens, Nicky; Van der Voort, Peter; Van Der Werf, Sylvie; van Dyk, Marlice; van Gulik, Laura; Van Hattem, Jarne; van Lelyveld, Steven; van Netten, Carolien; Van Twillert, Gitte; van Veen, Ilonka; Vanel, Noémie; Vanoverschelde, Henk; Vasudayan, Shoban Raj; Vauchy, Charline; Veeran, Shaminee; Veislinger, Aurélie; Ventura, Sara; Verbon, Annelies; Vidal, José Ernesto; Vieira, César; Vijayan, Deepak; Villanueva, Joy Ann; Villar, Judit; Villeneuve, Pierre-Marc; Villoldo, Andrea; Vinh Chau, Nguyen Van; Vishwanathan, Gayatri; Visseaux, Benoit; Visser, Hannah; Vitiello, Chiara; Vonkeman, Harald; Vuotto, Fanny; Wahab, Suhaila Abdul; Wahid, Nadirah Abdul; Wainstein, Marina; Wang, Chih-Hsien; Wei, Jia; Weil, Katharina; Wen, Tan Pei; Wesselius, Sanne; West, T. Eoin; Wham, Murray; Whelan, Bryan; White, Nicole; Wicky, Paul Henri; Wiedemann, Aurélie; Wijaya, Surya Otto; Wille, Keith; Willems, Suzette; Williams, Virginie; Wils, Evert-Jan; Wong, Calvin; Wong, Xin Ci; Wong, Yew Sing; Wong, Teck Fung; Xian, Gan Ee; Xian, Lim Saio; Xuan, Kuan Pei; Xynogalas, Ioannis; Yacoub, Sophie; Yakop, Siti Rohani Binti Mohd; Yamazaki, Masaki; Yazdanpanah, Yazdan; Yee Liang Hing, Nicholas; Yelnik, Cécile; Yeoh, Chian Hui; Yerkovich, Stephanie; Yokoyama, Toshiki; Yonis, Hodane; Yuliarto, Saptadi; Zaaqoq, Akram; Zabbe, Marion; Zacharowski, Kai; Zahid, Masliza; Zahran, Maram; Zaidan, Nor Zaila Binti; Zambon, Maria; Zambrano, Miguel; Zanella, Alberto; Zawadka, Konrad; Zaynah, Nurul; Zayyad, Hiba; Zoufaly, Alexander; Zucman, David.

