## Supplementary material for "Characteristics and outcomes of an international cohort of 400,000 hospitalised patients with Covid-19"

**Supplementary figure 1: Numbers of patients by country**

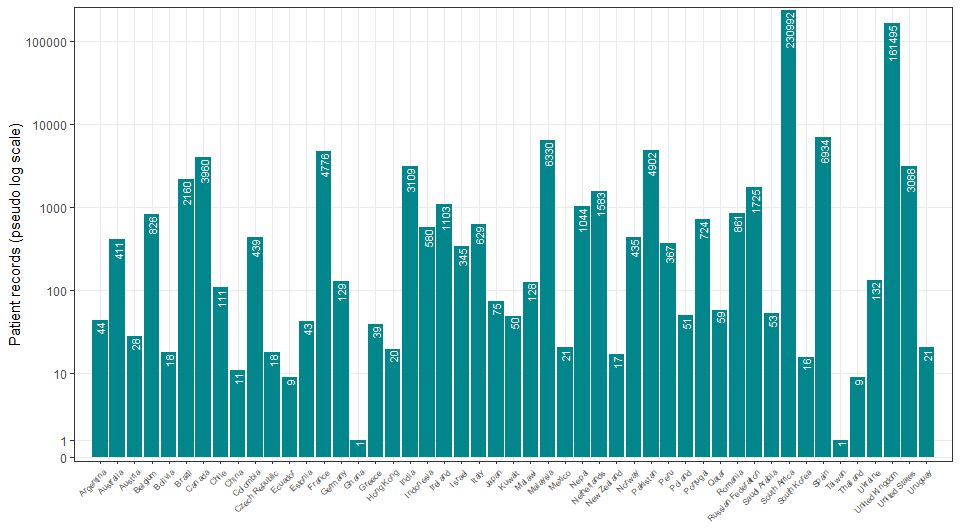

**Supplementary table 1: Participants’ characteristics by admission to ICU**

| Admission to ICU | Total N |  | Direct admission | Later admission | Never | (Missing) | Total |
| --- | --- | --- | --- | --- | --- | --- | --- |
| Total N (%) |  |  | 21890 (5.0) | 34375 (7.8) | 361614 (82.2) | 22043 (5.0) | 439922 |
| Age | 417879 (100.0) | Median (IQR) | 60.0 (50.0 to 69.0) | 60.0 (49.0 to 70.0) | 59.0 (44.0 to 75.0) | 61.0 (49.0 to 75.0) | 60.0 (45.0 to 74.0) |
|  | 417879 (100.0) | 0-9 | 192 (0.9) | 185 (0.5) | 4612 (1.3) | 282 (1.3) | 5271 (1.2) |
|  |  | 10-19 | 181 (0.8) | 243 (0.7) | 6649 (1.8) | 236 (1.1) | 7309 (1.7) |
|  |  | 20-29 | 584 (2.7) | 1012 (2.9) | 20396 (5.6) | 690 (3.1) | 22682 (5.2) |
|  |  | 30-39 | 1584 (7.2) | 2625 (7.6) | 37842 (10.5) | 1722 (7.8) | 43773 (10.0) |
|  |  | 40-49 | 2904 (13.3) | 4727 (13.8) | 47148 (13.0) | 2849 (12.9) | 57628 (13.1) |
|  |  | 50-59 | 5197 (23.7) | 7975 (23.2) | 64315 (17.8) | 4391 (19.9) | 81878 (18.6) |
|  |  | 60-69 | 5848 (26.7) | 8517 (24.8) | 60330 (16.7) | 4326 (19.6) | 79021 (18.0) |
|  |  | 70-79 | 4000 (18.3) | 6172 (18.0) | 55578 (15.4) | 3783 (17.2) | 69533 (15.8) |
|  |  | 80-89 | 1235 (5.6) | 2485 (7.2) | 48671 (13.5) | 2879 (13.1) | 55270 (12.6) |
|  |  | 90+ | 165 (0.8) | 434 (1.3) | 16073 (4.4) | 885 (4.0) | 17557 (4.0) |
| Region | 417879 (100.0) | East Asia & Pacific | 559 (2.6) | 724 (2.1) | 6079 (1.7) | 108 (0.5) | 7470 (1.7) |
|  |  | Europe & Central Asia | 9904 (45.2) | 19842 (57.7) | 142545 (39.4) | 9240 (41.9) | 181531 (41.3) |
|  |  | Latin America & Caribbean | 1514 (6.9) | 542 (1.6) | 1051 (0.3) | 83 (0.4) | 3190 (0.7) |
|  |  | Middle East & North Africa | 36 (0.2) | 106 (0.3) | 320 (0.1) | 45 (0.2) | 507 (0.1) |
|  |  | North America | 1615 (7.4) | 1798 (5.2) | 3168 (0.9) | 467 (2.1) | 7048 (1.6) |
|  |  | South Asia | 6687 (30.5) | 967 (2.8) | 1283 (0.4) | 118 (0.5) | 9055 (2.1) |
|  |  | Sub-Saharan Africa | 1575 (7.2) | 10396 (30.2) | 207168 (57.3) | 11982 (54.4) | 231121 (52.5) |
| Onset to admission (days) | 247283 (59.2) | Median (IQR) | 6.0 (2.0 to 9.0) | 3.0 (0.0 to 7.0) | 2.0 (0.0 to 7.0) | 2.0 (0.0 to 8.0) | 2.0 (0.0 to 7.0) |
| Length of hospital stay (days) | 417879 (100.0) | Median (IQR) | 9.0 (3.0 to 19.0) | 13.0 (6.0 to 26.0) | 5.0 (0.0 to 15.0) | 0.0 (0.0 to 12.0) | 6.0 (1.0 to 16.0) |
| Body mass index (BMI) (kg/m²) | 13397 (3.2) | Median (IQR) | 28.1 (24.7 to 32.3) | 27.8 (24.7 to 32.4) | 27.4 (23.5 to 31.2) | 27.9 (25.1 to 31.9) | 27.7 (24.2 to 31.8) |
| Heart rate on admission (bpm) | 168830 (40.4) | Median (IQR) | 94.0 (83.0 to 107.0) | 92.0 (81.0 to 105.0) | 89.0 (77.0 to 102.0) | 90.0 (78.0 to 103.0) | 90.0 (78.0 to 103.0) |
| Systolic blood pressure on admission (mmHg) | 175288 (41.9) | Median (IQR) | 130.0 (116.0 to 142.0) | 128.0 (116.0 to 142.0) | 130.0 (116.0 to 145.0) | 129.0 (115.0 to 145.0) | 130.0 (116.0 to 144.0) |
| Diastolic blood pressure on admission (mmHg) | 175336 (42.0) | Median (IQR) | 74.0 (65.0 to 82.0) | 74.0 (67.0 to 82.0) | 75.0 (66.0 to 84.0) | 74.0 (65.0 to 82.0) | 74.0 (66.0 to 83.0) |
| Temperature on admission (degrees C) | 176527 (42.2) | Median (IQR) | 37.0 (36.7 to 37.8) | 37.2 (36.5 to 38.1) | 37.0 (36.5 to 37.8) | 37.0 (36.5 to 37.8) | 37.0 (36.5 to 37.8) |
| Oxygen saturation on admission (%) | 177315 (42.4) | Median (IQR) | 93.0 (89.0 to 96.0) | 94.0 (91.0 to 96.0) | 96.0 (94.0 to 98.0) | 96.0 (93.0 to 97.0) | 96.0 (93.0 to 97.0) |
| - On oxygen therapy | 55532 (13.3) | Median (IQR) | 93.0 (89.0 to 96.0) | 95.0 (92.0 to 97.0) | 95.0 (93.0 to 97.0) | 95.0 (92.0 to 97.0) | 95.0 (92.0 to 97.0) |
| - In room air | 113124 (27.1) | Median (IQR) | 94.0 (88.0 to 97.0) | 94.0 (89.0 to 96.0) | 96.0 (94.0 to 98.0) | 96.0 (94.0 to 98.0) | 96.0 (93.0 to 98.0) |
| - Unknown oxygenation status | 8828 (2.1) | Median (IQR) | 94.0 (91.0 to 97.0) | 95.0 (91.0 to 97.0) | 96.0 (93.0 to 98.0) | 96.0 (93.0 to 97.0) | 96.0 (93.0 to 98.0) |
| Respiratory rate on admission | 169114 (40.5) | Median (IQR) | 24.0 (20.0 to 30.0) | 22.0 (20.0 to 28.0) | 20.0 (18.0 to 24.0) | 20.0 (18.0 to 25.0) | 20.0 (18.0 to 25.0) |
| Outcome | 417879 (100.0) | Censored | 5136 (23.5) | 3252 (9.5) | 18376 (5.1) | 4017 (18.2) | 30781 (7.0) |
|  |  | Death | 7631 (34.9) | 11072 (32.2) | 77632 (21.5) | 5846 (26.5) | 102181 (23.2) |
|  |  | Discharge | 9123 (41.7) | 20051 (58.3) | 265606 (73.5) | 12180 (55.3) | 306960 (69.8) |
| SARS-CoV-2 status | 417879 (100.0) | Positive | 20287 (92.7) | 32987 (96.0) | 329753 (91.2) | 20405 (92.6) | 403432 (91.7) |
|  |  | Unknown | 1603 (7.3) | 1388 (4.0) | 31861 (8.8) | 1638 (7.4) | 36490 (8.3) |

**Supplementary table 2: Participants’ characteristics by age**

| Age | Total N |  | [0,10) | [10,20) | [20,50) | [50,80) | [80,120] | Total |
| --- | --- | --- | --- | --- | --- | --- | --- | --- |
| Total N (%) |  |  | 5271 (1.2) | 7309 (1.7) | 124083 (28.2) | 230432 (52.4) | 72827 (16.6) | 439922 |
| Body mass index (BMI) (kg/m²) | 13949 (3.2) | Median (IQR) | 15.9 (13.0 to 17.7) | 23.6 (19.6 to 29.5) | 28.5 (24.8 to 33.0) | 27.8 (24.5 to 31.8) | 25.7 (21.9 to 29.3) | 27.7 (24.2 to 31.8) |
| Height (cm) | 13983 (3.2) | Median (IQR) | 99.0 (63.0 to 154.9) | 165.0 (158.5 to 173.0) | 169.2 (163.0 to 175.3) | 168.0 (161.0 to 175.0) | 165.0 (158.0 to 171.0) | 168.0 (160.0 to 175.0) |
| Heart rate on admission (bpm) | 174714 (39.7) | Median (IQR) | 138.0 (115.0 to 157.0) | 100.0 (86.0 to 114.0) | 97.0 (85.0 to 109.0) | 90.0 (79.0 to 102.0) | 85.0 (74.0 to 97.0) | 90.0 (78.0 to 103.0) |
| Systolic blood pressure on admission (mmHg) | 181293 (41.2) | Median (IQR) | 101.0 (89.0 to 113.0) | 119.0 (110.0 to 129.0) | 125.0 (115.0 to 137.0) | 130.0 (116.0 to 145.0) | 133.0 (117.0 to 150.0) | 130.0 (116.0 to 144.0) |
| Diastolic blood pressure on admission (mmHg) | 181361 (41.2) | Median (IQR) | 60.0 (49.0 to 70.0) | 72.0 (64.0 to 80.0) | 77.0 (70.0 to 85.0) | 75.0 (67.0 to 83.0) | 72.0 (63.0 to 82.0) | 74.0 (66.0 to 83.0) |
| Temperature on admission (degrees C) | 182514 (41.5) | Median (IQR) | 37.0 (36.6 to 37.9) | 36.9 (36.5 to 37.3) | 37.0 (36.6 to 37.9) | 37.1 (36.6 to 37.9) | 36.9 (36.4 to 37.6) | 37.0 (36.5 to 37.8) |
| Oxygen saturation on admission (%) | 183304 (41.7) | Median (IQR) | 98.0 (97.0 to 100.0) | 98.0 (97.0 to 99.0) | 97.0 (94.0 to 98.0) | 95.0 (92.0 to 97.0) | 96.0 (93.0 to 97.0) | 96.0 (93.0 to 97.0) |
| - On oxygen therapy | 57687 (13.1) | Median (IQR) | 97.0 (94.0 to 99.0) | 97.0 (94.0 to 99.0) | 95.0 (93.0 to 97.0) | 95.0 (92.0 to 97.0) | 95.0 (92.0 to 97.0) | 95.0 (92.0 to 97.0) |
| - In room air | 116762 (26.5) | Median (IQR) | 98.0 (97.0 to 100.0) | 98.0 (97.0 to 99.0) | 97.0 (95.0 to 98.0) | 95.0 (92.0 to 97.0) | 96.0 (94.0 to 97.0) | 96.0 (93.0 to 98.0) |
| - Unknown oxygenation status | 9042 (2.1) | Median (IQR) | 98.0 (96.0 to 100.0) | 98.0 (96.0 to 99.0) | 96.0 (94.0 to 98.0) | 95.0 (92.0 to 97.0) | 96.0 (93.0 to 97.0) | 96.0 (93.0 to 98.0) |
| Respiratory rate on admission | 175080 (39.8) | Median (IQR) | 33.0 (25.0 to 42.0) | 20.0 (18.0 to 22.0) | 20.0 (18.0 to 24.0) | 21.0 (18.0 to 25.0) | 20.0 (18.0 to 24.0) | 20.0 (18.0 to 25.0) |
| Admission to ICU | 417879 (95.0) | Never | 4612 (92.4) | 6649 (94.0) | 105386 (88.7) | 180223 (82.7) | 64744 (93.7) | 361614 (86.5) |
|  |  | Later admission | 185 (3.7) | 243 (3.4) | 8364 (7.0) | 22664 (10.4) | 2919 (4.2) | 34375 (8.2) |
|  |  | Direct admission | 192 (3.8) | 181 (2.6) | 5072 (4.3) | 15045 (6.9) | 1400 (2.0) | 21890 (5.2) |
| Ever received invasive mechanical ventilation | 431574 (98.1) | No | 4961 (95.1) | 6994 (96.4) | 115178 (94.0) | 202674 (89.8) | 69244 (97.8) | 399051 (92.5) |
|  |  | Yes | 254 (4.9) | 262 (3.6) | 7343 (6.0) | 23099 (10.2) | 1565 (2.2) | 32523 (7.5) |
| Ever received non-invasive ventilation | 299309 (68.0) | No | 3315 (95.9) | 5098 (97.8) | 67113 (92.7) | 132339 (85.5) | 59185 (93.3) | 267050 (89.2) |
|  |  | Yes | 140 (4.1) | 113 (2.2) | 5276 (7.3) | 22511 (14.5) | 4219 (6.7) | 32259 (10.8) |
| High flow nasal cannula | 276536 (62.9) | No | 2970 (93.7) | 4290 (96.0) | 57376 (88.5) | 116110 (80.6) | 49960 (83.3) | 230706 (83.4) |
|  |  | Yes | 199 (6.3) | 181 (4.0) | 7431 (11.5) | 27985 (19.4) | 10034 (16.7) | 45830 (16.6) |
| Oxygen therapy via mask | 58 (0.0) | No | 0 (NaN) | 0 (NaN) | 5 (100.0) | 35 (85.4) | 11 (91.7) | 51 (87.9) |
|  |  | Yes | 0 (NaN) | 0 (NaN) | 0 (0.0) | 6 (14.6) | 1 (8.3) | 7 (12.1) |
| Ever received any oxygen supplementation | 435429 (99.0) | No | 4215 (80.5) | 6046 (83.1) | 78644 (63.8) | 98846 (43.4) | 28895 (40.4) | 216646 (49.8) |
|  |  | Yes | 1022 (19.5) | 1231 (16.9) | 44688 (36.2) | 129158 (56.6) | 42684 (59.6) | 218783 (50.2) |
| Outcome | 439922 (100.0) | Censored | 318 (6.0) | 516 (7.1) | 6815 (5.5) | 16579 (7.2) | 6553 (9.0) | 30781 (7.0) |
|  |  | Death | 168 (3.2) | 223 (3.1) | 11093 (8.9) | 61387 (26.6) | 29310 (40.2) | 102181 (23.2) |
|  |  | Discharge | 4785 (90.8) | 6570 (89.9) | 106175 (85.6) | 152466 (66.2) | 36964 (50.8) | 306960 (69.8) |
| SARS-CoV-2 status | 439922 (100.0) | Positive | 4494 (85.3) | 6059 (82.9) | 111256 (89.7) | 211349 (91.7) | 70274 (96.5) | 403432 (91.7) |
|  |  | Unknown | 777 (14.7) | 1250 (17.1) | 12827 (10.3) | 19083 (8.3) | 2553 (3.5) | 36490 (8.3) |

**Supplementary table 3: Symptom prevalence**

| Symptom | N recorded (%) |  | N (%) |
| --- | --- | --- | --- |
| Abdominal pain | 168925 (80.8) | No | 154387 (73.9) |
|  |  | Yes | 14538 (7.0) |
|  |  | (Missing) | 40027 (19.2) |
| Altered consciousness/confusion | 168647 (80.7) | No | 134495 (64.4) |
|  |  | Yes | 34152 (16.3) |
|  |  | (Missing) | 40305 (19.3) |
| Anorexia | 18220 (8.7) | No | 16202 (7.8) |
|  |  | Yes | 2018 (1.0) |
|  |  | (Missing) | 190732 (91.3) |
| Asymptomatic | 108118 (51.7) | No | 98501 (47.1) |
|  |  | Yes | 9617 (4.6) |
|  |  | (Missing) | 100834 (48.3) |
| Bleeding | 167620 (80.2) | No | 164676 (78.8) |
|  |  | Yes | 2944 (1.4) |
|  |  | (Missing) | 41332 (19.8) |
| Chest pain | 170076 (81.4) | No | 149052 (71.3) |
|  |  | Yes | 21024 (10.1) |
|  |  | (Missing) | 38876 (18.6) |
| Conjunctivitis | 160342 (76.7) | No | 159749 (76.5) |
|  |  | Yes | 593 (0.3) |
|  |  | (Missing) | 48610 (23.3) |
| Cough | 183949 (88.0) | No | 80990 (38.8) |
|  |  | Yes | 102959 (49.3) |
|  |  | (Missing) | 25003 (12.0) |
| Diarrhoea | 173097 (82.8) | No | 146380 (70.1) |
|  |  | Yes | 26717 (12.8) |
|  |  | (Missing) | 35855 (17.2) |
| Ear pain | 138710 (66.4) | No | 138215 (66.1) |
|  |  | Yes | 495 (0.2) |
|  |  | (Missing) | 70242 (33.6) |
| Fatigue/malaise | 166624 (79.7) | No | 103763 (49.7) |
|  |  | Yes | 62861 (30.1) |
|  |  | (Missing) | 42328 (20.3) |
| Headache | 160520 (76.8) | No | 144113 (69.0) |
|  |  | Yes | 16407 (7.9) |
|  |  | (Missing) | 48432 (23.2) |
| Fever | 184093 (88.1) | No | 86366 (41.3) |
|  |  | Yes | 97727 (46.8) |
|  |  | (Missing) | 24859 (11.9) |
| Inability to walk | 23325 (11.2) | No | 22179 (10.6) |
|  |  | Yes | 1146 (0.5) |
|  |  | (Missing) | 185627 (88.8) |
| Lost/altered sense of smell | 127330 (60.9) | No | 119605 (57.2) |
|  |  | Yes | 7725 (3.7) |
|  |  | (Missing) | 81622 (39.1) |
| Lost/altered sense of taste | 124407 (59.5) | No | 115286 (55.2) |
|  |  | Yes | 9121 (4.4) |
|  |  | (Missing) | 84545 (40.5) |
| Lymphadenopathy | 153539 (73.5) | No | 152758 (73.1) |
|  |  | Yes | 781 (0.4) |
|  |  | (Missing) | 55413 (26.5) |
| Muscle ache/joint pain | 160593 (76.9) | No | 133269 (63.8) |
|  |  | Yes | 27324 (13.1) |
|  |  | (Missing) | 48359 (23.1) |
| Runny nose | 155193 (74.3) | No | 150386 (72.0) |
|  |  | Yes | 4807 (2.3) |
|  |  | (Missing) | 53759 (25.7) |
| Seizures | 162921 (78.0) | No | 161012 (77.1) |
|  |  | Yes | 1909 (0.9) |
|  |  | (Missing) | 46031 (22.0) |
| Severe dehydration | 78295 (37.5) | No | 68827 (32.9) |
|  |  | Yes | 9468 (4.5) |
|  |  | (Missing) | 130657 (62.5) |
| Shortness of breath | 184451 (88.3) | No | 77885 (37.3) |
|  |  | Yes | 106566 (51.0) |
|  |  | (Missing) | 24501 (11.7) |
| Skin rash | 162711 (77.9) | No | 158612 (75.9) |
|  |  | Yes | 4099 (2.0) |
|  |  | (Missing) | 46241 (22.1) |
| Sore throat | 155945 (74.6) | No | 145154 (69.5) |
|  |  | Yes | 10791 (5.2) |
|  |  | (Missing) | 53007 (25.4) |
| Vomiting/nausea | 172613 (82.6) | No | 144702 (69.3) |
|  |  | Yes | 27911 (13.4) |
|  |  | (Missing) | 36339 (17.4) |
| Wheezing | 163231 (78.1) | No | 153689 (73.6) |
|  |  | Yes | 9542 (4.6) |
|  |  | (Missing) | 45721 (21.9) |
| WHO | 105863 (50.7) | Yes | 105863 (50.7) |
|  |  | (Missing) | 103089 (49.3) |
| CDC | 136515 (65.3) | Yes | 136515 (65.3) |
|  |  | (Missing) | 72437 (34.7) |
| PHE | 131630 (63.0) | Yes | 131630 (63.0) |
|  |  | (Missing) | 77322 (37.0) |
| ECDC | 147604 (70.6) | Yes | 147604 (70.6) |
|  |  | (Missing) | 61348 (29.4) |

**Supplementary figure 2: Proportions of symptom prevalence by age for each country** Countries with fewer than 500 are grouped into ‘other’

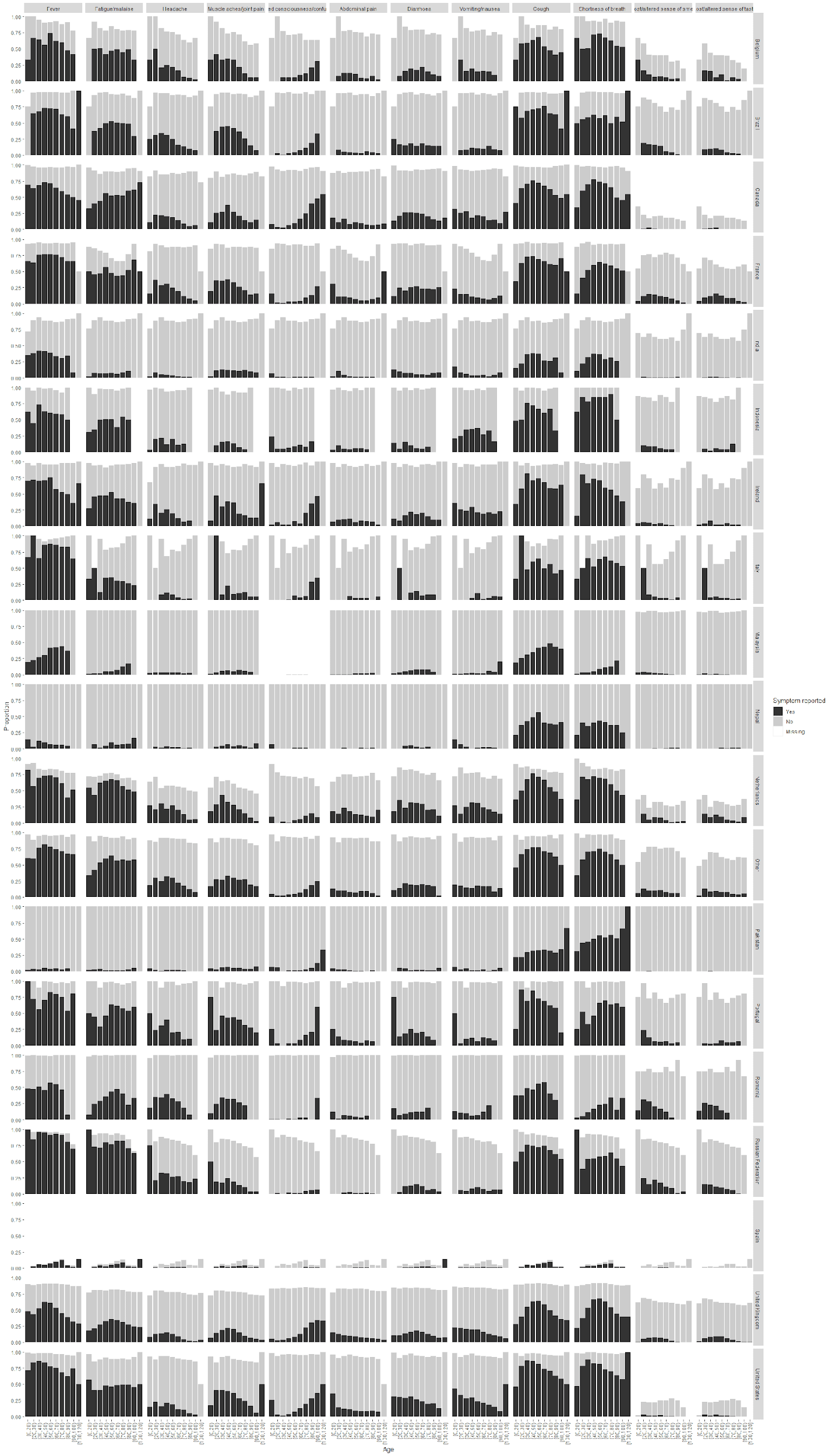

**Supplementary figure 3: Proportions of symptom prevalence by age among individuals with confirmed SARS-CoV-2**

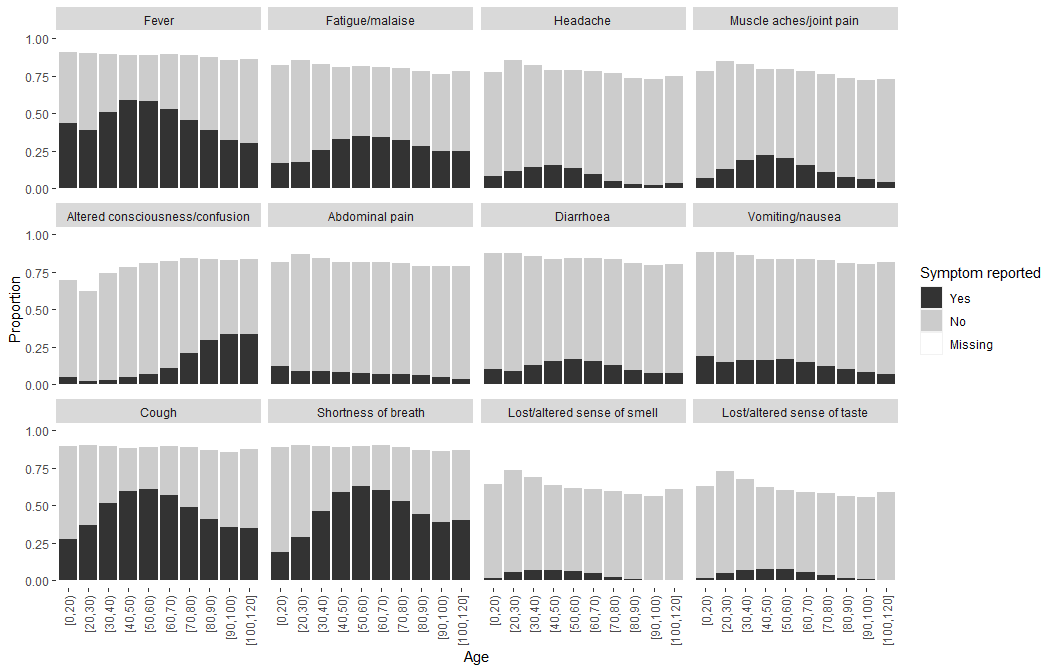

**Supplementary figure 4: Proportions of symptom prevalence by age among individuals without confirmed SARS-CoV-2**

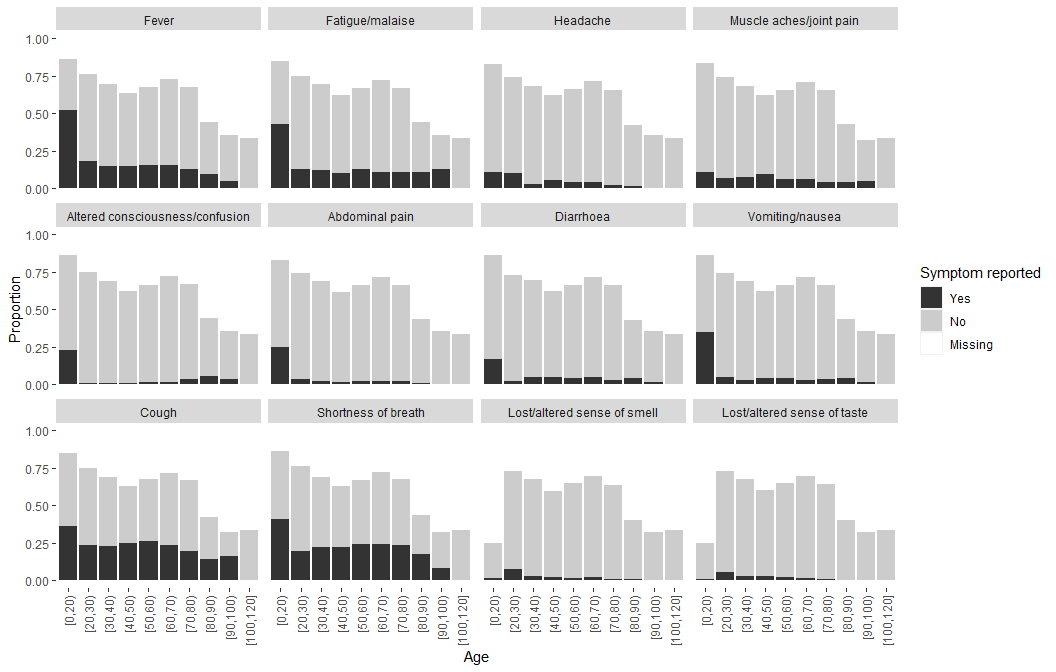

**Supplementary figure 5: Proportions with each symptom by SARS-CoV-2 status**

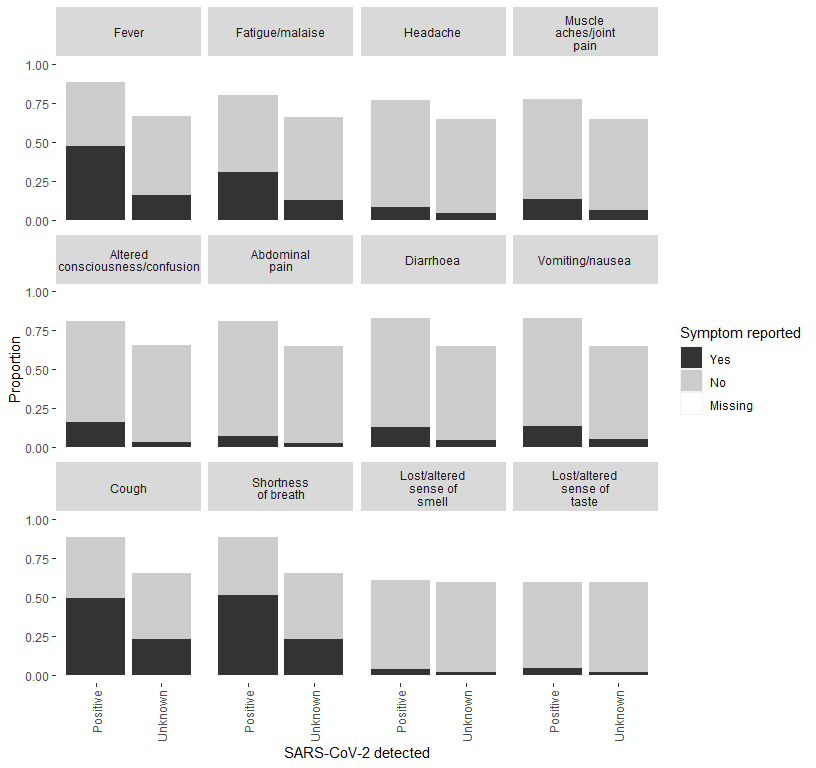

**Supplementary figure 6: Proportions meeting each symptom definition by SARS-CoV-2 status**

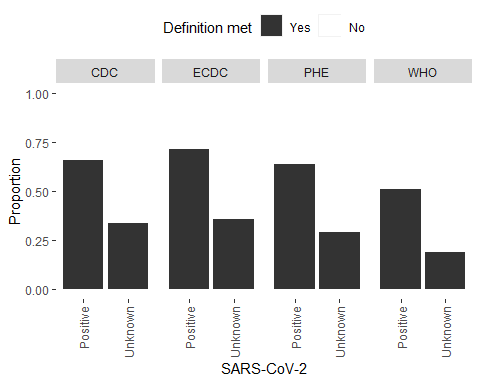

**Supplementary figure 7: Proportions meeting each symptom definition by age and SARS-CoV-2 status**

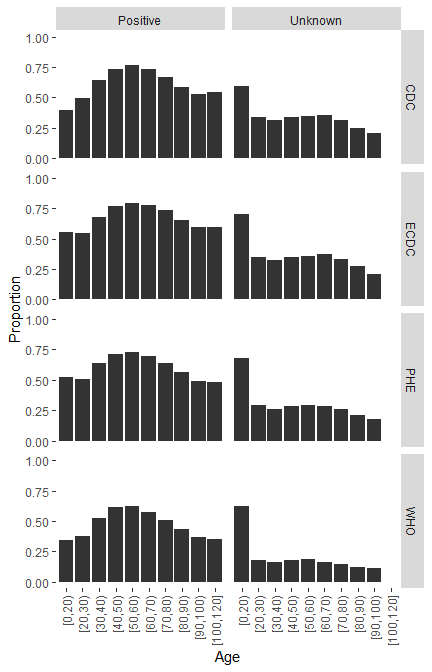

**Supplementary table 4: Routine laboratory test results.** Patients with lymphocyte count plus neutrophil count greater than total white blood cell count were excluded

|  | N | Mean (SD) | Median (IQR) |
| --- | --- | --- | --- |
| **Inflammation** |  |  |  |
| C-Reactive Protein (mg/L) | 83569 (83.0) | 94.7 (76.3) | 76.0 (32.0 to 140.0) |
| White blood cell count ($\times{10}^{9}$ cells per L) | 100710 (100.0) | 8.0 (3.5) | 7.3 (5.5 to 9.8) |
| Neutrophil count ($\times{10}^{9}$ cells per L) | 100710 (100.0) | 6.3 (3.3) | 5.5 (3.9 to 7.9) |
| Lymphocyte count ($\times{10}^{9}$ cells per L) | 100710 (100.0) | 1.1 (0.7) | 0.9 (0.6 to 1.3) |
| **Liver function tests** |  |  |  |
| Alanine aminotransferase (U/L) | 61973 (61.5) | 34.6 (25.1) | 27.0 (17.0 to 43.0) |
| Aspartate aminotransferase (U/L) | 12236 (12.1) | 45.3 (31.5) | 34.0 (23.1 to 57.0) |
| Total bilirubin ($\mu$mol/L) | 66082 (65.6) | 10.7 (6.0) | 9.0 (7.0 to 13.0) |
| **Kidney function tests** |  |  |  |
| Urea (mmol/L) | 87348 (86.7) | 8.4 (5.9) | 6.5 (4.6 to 10.1) |

**Supplementary figure 8: Routine laboratory test values by age** Patients with lymphocyte count plus neutrophic count greater than total white blood cell count were excluded

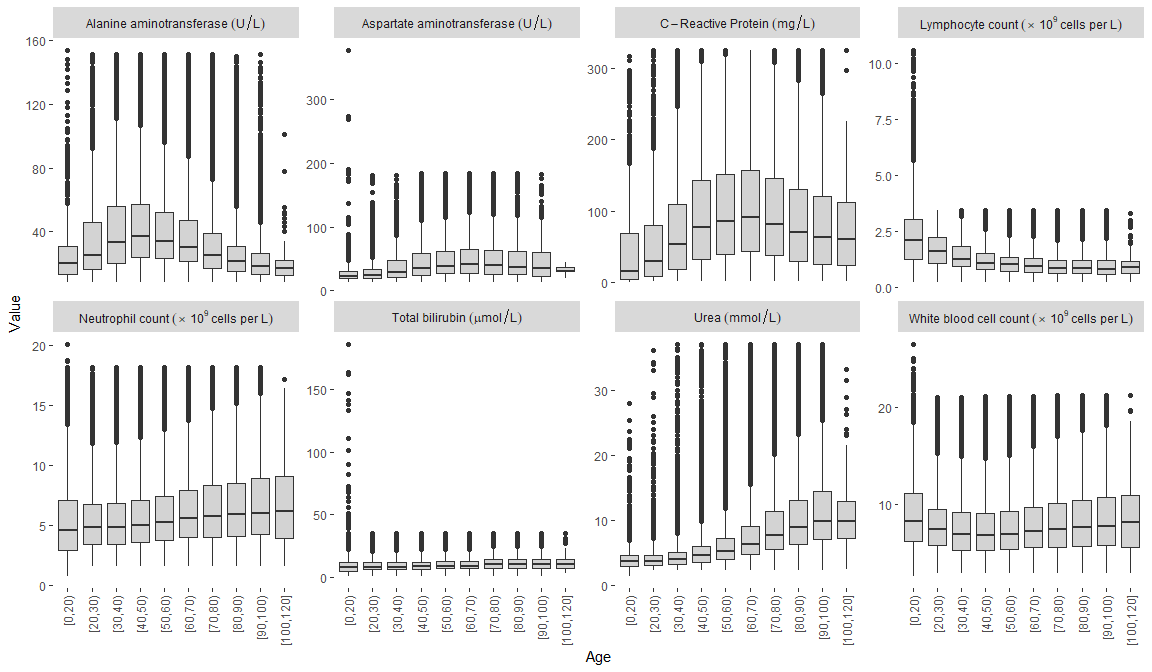

**Supplementary table 5: Proportions of individuals with pre-existing comorbidities and risk factors**

| Comorbidity/risk factor | Total N |  | N (%) |
| --- | --- | --- | --- |
| HIV | 348537 (79.2) | No | 332156 (75.5) |
|  |  | Yes | 16381 (3.7) |
|  |  | (Missing) | 91385 (20.8) |
| Asthma | 355800 (80.9) | No | 323331 (73.5) |
|  |  | Yes | 32469 (7.4) |
|  |  | (Missing) | 84122 (19.1) |
| Chronic cardiac disease | 351514 (79.9) | No | 294247 (66.9) |
|  |  | Yes | 57267 (13.0) |
|  |  | (Missing) | 88408 (20.1) |
| Chronic hematologic disease | 185813 (42.2) | No | 178813 (40.6) |
|  |  | Yes | 7000 (1.6) |
|  |  | (Missing) | 254109 (57.8) |
| Chronic kidney disease | 352463 (80.1) | No | 320185 (72.8) |
|  |  | Yes | 32278 (7.3) |
|  |  | (Missing) | 87459 (19.9) |
| Chronic neurological disorder | 191845 (43.6) | No | 171303 (38.9) |
|  |  | Yes | 20542 (4.7) |
|  |  | (Missing) | 248077 (56.4) |
| Chronic pulmonary disease | 356128 (81.0) | No | 322055 (73.2) |
|  |  | Yes | 34073 (7.7) |
|  |  | (Missing) | 83794 (19.0) |
| Dementia | 186579 (42.4) | No | 165575 (37.6) |
|  |  | Yes | 21004 (4.8) |
|  |  | (Missing) | 253343 (57.6) |
| Diabetes | 367518 (83.5) | No | 263208 (59.8) |
|  |  | Yes | 104310 (23.7) |
|  |  | (Missing) | 72404 (16.5) |
| Hypertension | 343503 (78.1) | No | 201756 (45.9) |
|  |  | Yes | 141747 (32.2) |
|  |  | (Missing) | 96419 (21.9) |
| Immunosuppression | 117193 (26.6) | No | 113989 (25.9) |
|  |  | Yes | 3204 (0.7) |
|  |  | (Missing) | 322729 (73.4) |
| Liver disease | 194459 (44.2) | No | 188410 (42.8) |
|  |  | Yes | 6049 (1.4) |
|  |  | (Missing) | 245463 (55.8) |
| Malignant neoplasm | 354916 (80.7) | No | 334928 (76.1) |
|  |  | Yes | 19988 (4.5) |
|  |  | (Missing) | 85006 (19.3) |
| Malnutrition | 180778 (41.1) | No | 176703 (40.2) |
|  |  | Yes | 4075 (0.9) |
|  |  | (Missing) | 259144 (58.9) |
| Obesity | 236010 (53.6) | No | 202054 (45.9) |
|  |  | Yes | 33956 (7.7) |
|  |  | (Missing) | 203912 (46.4) |
| Other | 230970 (52.5) | No | 226267 (51.4) |
|  |  | Yes | 4703 (1.1) |
|  |  | (Missing) | 208952 (47.5) |
| Rare diseases and inborn errors of metabolism | 117439 (26.7) | No | 116962 (26.6) |
|  |  | Yes | 477 (0.1) |
|  |  | (Missing) | 322483 (73.3) |
| Rheumatologic disorder | 186003 (42.3) | No | 166497 (37.8) |
|  |  | Yes | 19506 (4.4) |
|  |  | (Missing) | 253919 (57.7) |
| Smoking | 151803 (34.5) | No | 101370 (23.0) |
|  |  | Yes | 50433 (11.5) |
|  |  | (Missing) | 288119 (65.5) |
| Transplantation | 118138 (26.9) | No | 117071 (26.6) |
|  |  | Yes | 1067 (0.2) |
|  |  | (Missing) | 321784 (73.1) |
| Tuberculosis | 194129 (44.1) | No | 187421 (42.6) |
|  |  | Yes | 6708 (1.5) |
|  |  | (Missing) | 245793 (55.9) |
| Pregnancy | 155936 (35.4) | No | 147381 (33.5) |
|  |  | Yes | 8555 (1.9) |
|  |  | (Missing) | 283986 (64.6) |

**Supplementary figure 9: Prevalence of pre-existing comorbidities and risk factors by age**

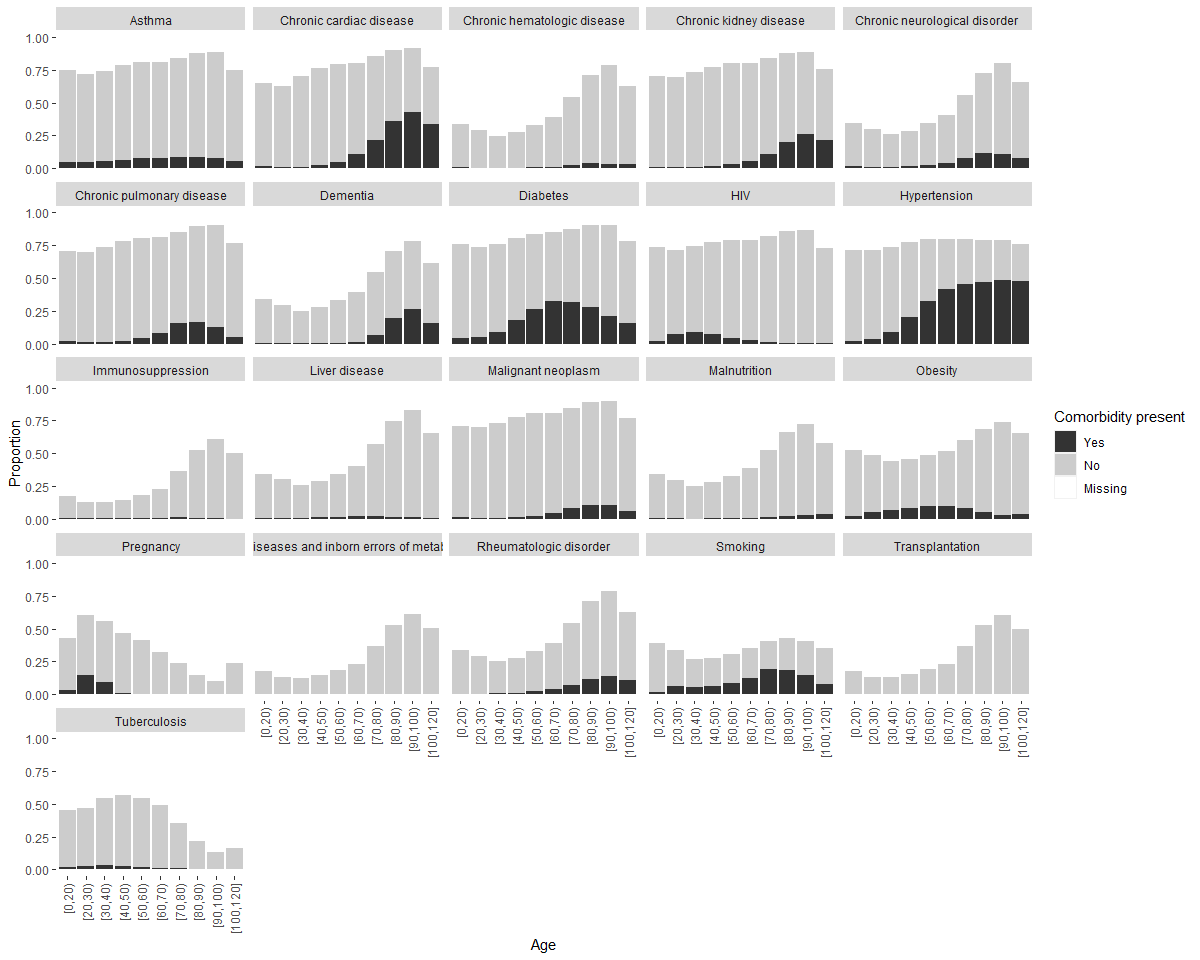

**Supplementary table 6: Number and proportion of patients receiving each treatment**

| Treatment | Total N |  | N (%) |
| --- | --- | --- | --- |
| Agents acting on the renin angiotensin system | 193560 (92.6) | No | 185602 (88.8) |
|  |  | Yes | 7958 (3.8) |
|  |  | (Missing) | 15392 (7.4) |
| Antiarrhythymic medications | 1489 (0.7) | No | 1403 (0.7) |
|  |  | Yes | 86 (0.0) |
|  |  | (Missing) | 207463 (99.3) |
| Antibacterial agents | 188595 (90.3) | No | 38695 (18.5) |
|  |  | Yes | 149900 (71.7) |
|  |  | (Missing) | 20357 (9.7) |
| Antifungal agents | 189806 (90.8) | No | 179593 (85.9) |
|  |  | Yes | 10213 (4.9) |
|  |  | (Missing) | 19146 (9.2) |
| Antimalarial agents | 9812 (4.7) | No | 5770 (2.8) |
|  |  | Yes | 4042 (1.9) |
|  |  | (Missing) | 199140 (95.3) |
| Antiviral agents | 183297 (87.7) | No | 151030 (72.3) |
|  |  | Yes | 32267 (15.4) |
|  |  | (Missing) | 25655 (12.3) |
| Convalescent plasma | 119654 (57.3) | No | 117157 (56.1) |
|  |  | Yes | 2497 (1.2) |
|  |  | (Missing) | 89298 (42.7) |
| Corticosteroids | 194033 (92.9) | No | 111225 (53.2) |
|  |  | Yes | 82808 (39.6) |
|  |  | (Missing) | 14919 (7.1) |
| Diuretics | 1911 (0.9) | No | 1698 (0.8) |
|  |  | Yes | 213 (0.1) |
|  |  | (Missing) | 207041 (99.1) |
| Experimental agents | 5865 (2.8) | No | 3884 (1.9) |
|  |  | Yes | 1981 (0.9) |
|  |  | (Missing) | 203087 (97.2) |
| Immunosuppressants | 8697 (4.2) | No | 6288 (3.0) |
|  |  | Yes | 2409 (1.2) |
|  |  | (Missing) | 200255 (95.8) |
| Inotropes/vasopressors | 191098 (91.5) | No | 175535 (84.0) |
|  |  | Yes | 15563 (7.4) |
|  |  | (Missing) | 17854 (8.5) |
| Interleukin inhibitors | 119239 (57.1) | No | 117355 (56.2) |
|  |  | Yes | 1884 (0.9) |
|  |  | (Missing) | 89713 (42.9) |
| Intravenous fluids | 5822 (2.8) | No | 905 (0.4) |
|  |  | Yes | 4917 (2.4) |
|  |  | (Missing) | 203130 (97.2) |
| Nasogastric fluids | 4299 (2.1) | No | 2118 (1.0) |
|  |  | Yes | 2181 (1.0) |
|  |  | (Missing) | 204653 (97.9) |
| Neuromuscular blocking agents | 45146 (21.6) | No | 39879 (19.1) |
|  |  | Yes | 5267 (2.5) |
|  |  | (Missing) | 163806 (78.4) |
| Non-steroidal anti-inflammatory agents | 5764 (2.8) | No | 4574 (2.2) |
|  |  | Yes | 1190 (0.6) |
|  |  | (Missing) | 203188 (97.2) |
| Off-label/compassionate use medications | 122917 (58.8) | No | 118293 (56.6) |
|  |  | Yes | 4624 (2.2) |
|  |  | (Missing) | 86035 (41.2) |
| Other interventions | 28444 (13.6) | No | 23938 (11.5) |
|  |  | Yes | 4506 (2.2) |
|  |  | (Missing) | 180508 (86.4) |
| Anticoagulant | 21215 (10.2) | No | 10173 (4.9) |
|  |  | Yes | 11042 (5.3) |
|  |  | (Missing) | 187737 (89.8) |
| Total parenteral nutrition | 6220 (3.0) | No | 6219 (3.0) |
|  |  | Yes | 1 (0.0) |
|  |  | (Missing) | 202732 (97.0) |
| Extracorporeal membrane oxygenation | 197946 (94.7) | No | 196642 (94.1) |
|  |  | Yes | 1304 (0.6) |
|  |  | (Missing) | 11006 (5.3) |

**Supplementary figure 10: Proportion receiving each treatment by age**

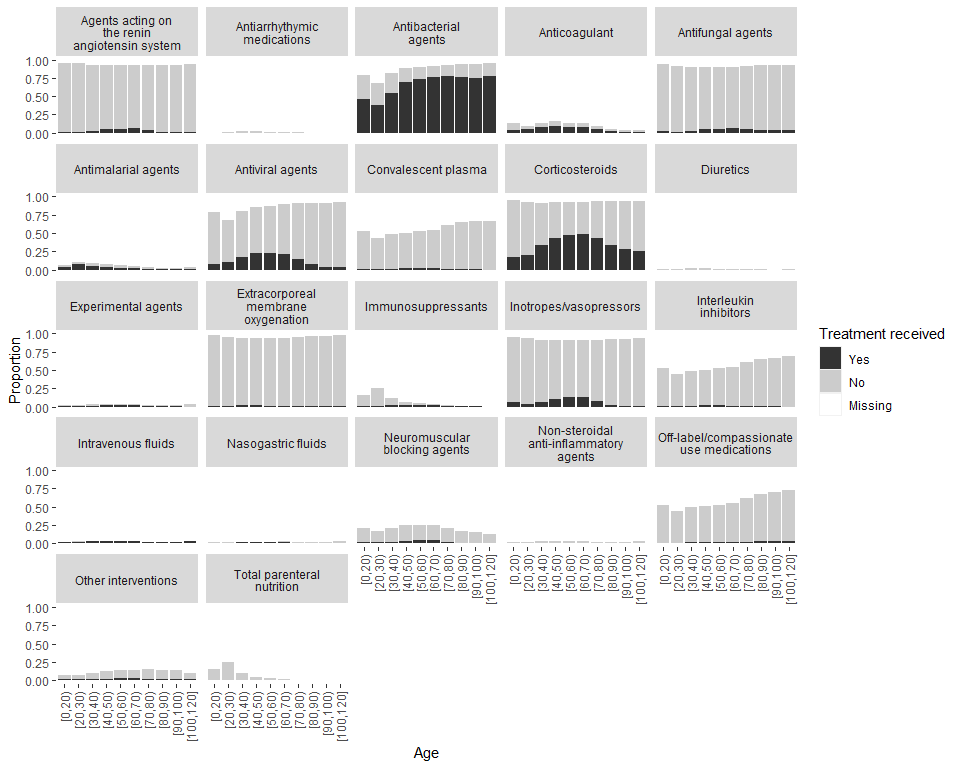

**Supplementary figure 11: Use of corticosteroids over time in all patients and in patients who did and did not receive oxygen therapy** The green line shows the month that results on corticosteroids from the RECOVERY trial were published.

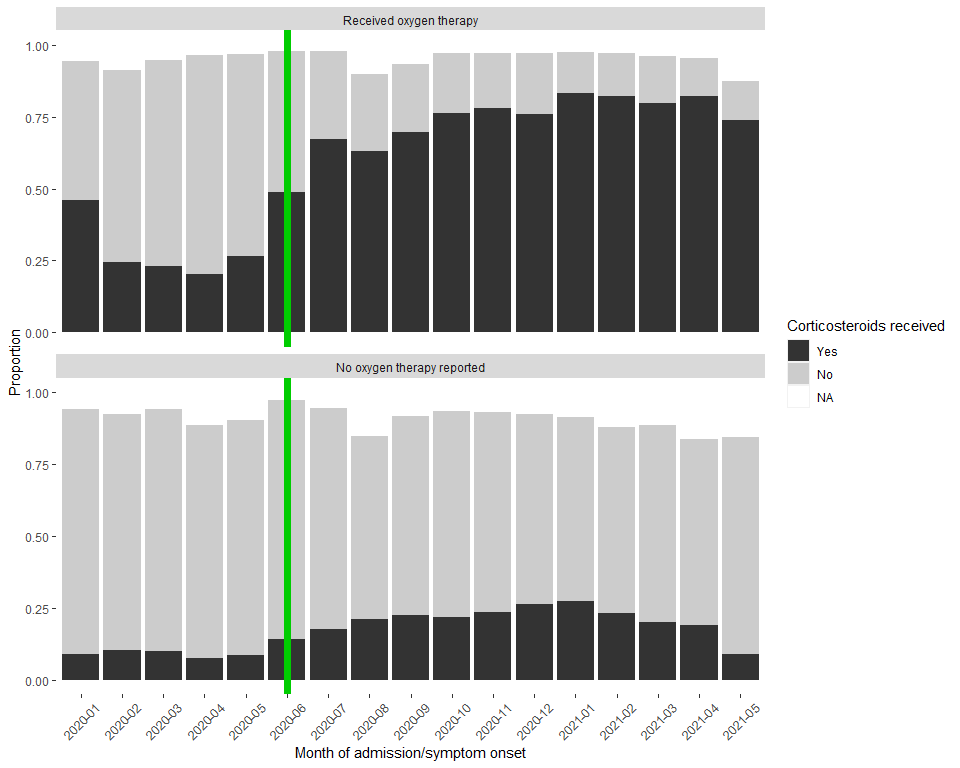

**Supplementary figure 12: CFR by month of admission in each country**

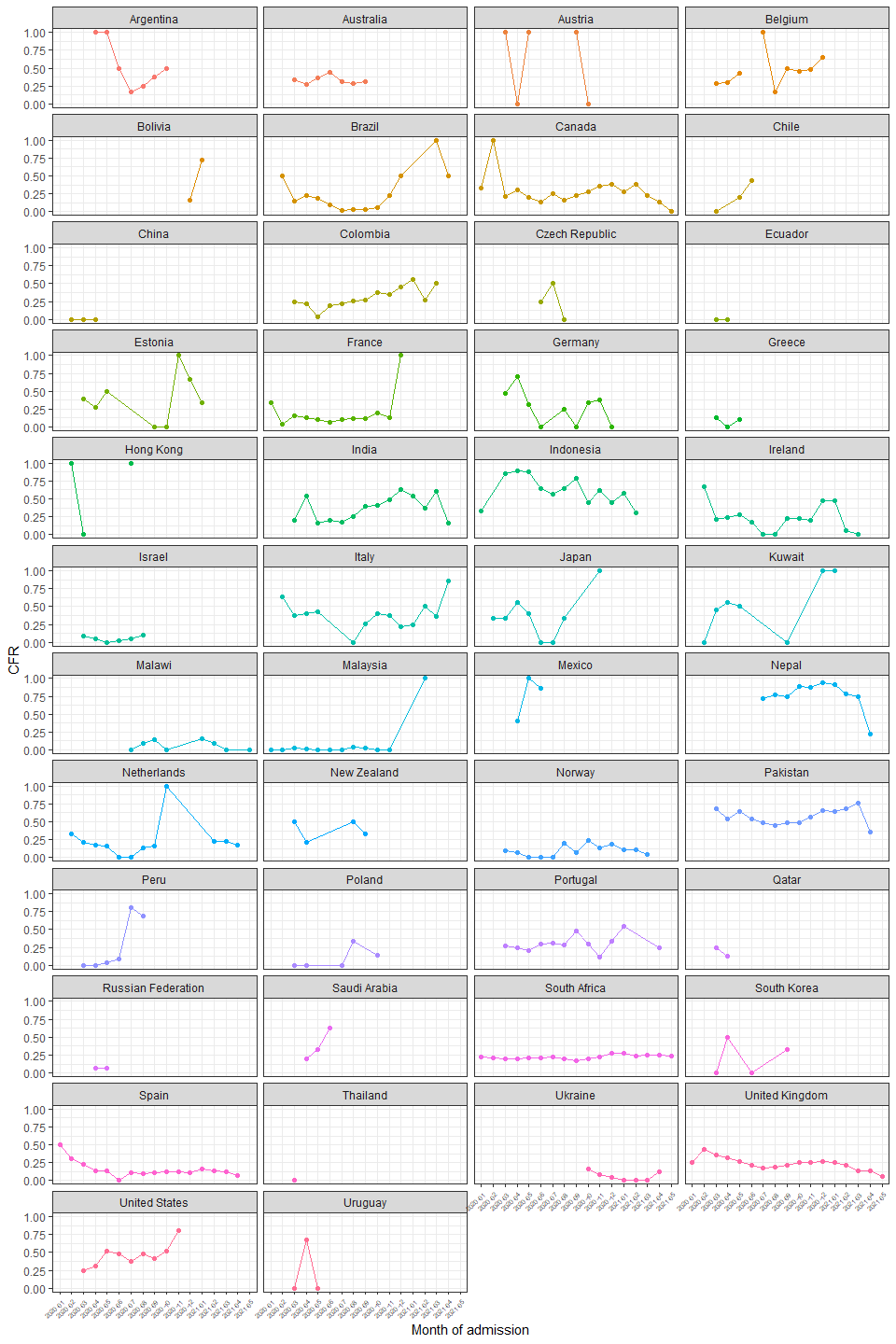

**Supplementary figure 13: Cumulative incidence curves of death and discharge**

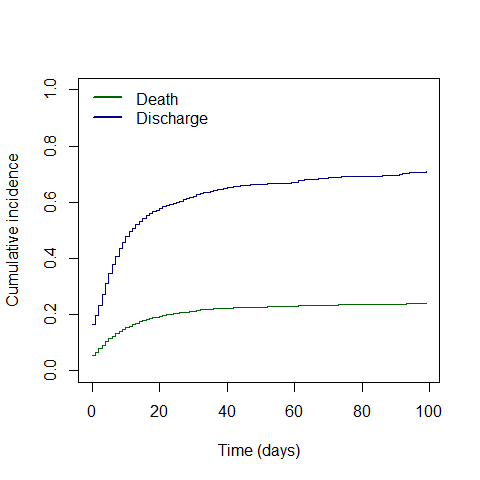

**Supplementary figure 14: Hazard ratios and 95% CIs for death by age group** Adjusted for sex and stratified by country; HRs are plotted on logarithmic scale.

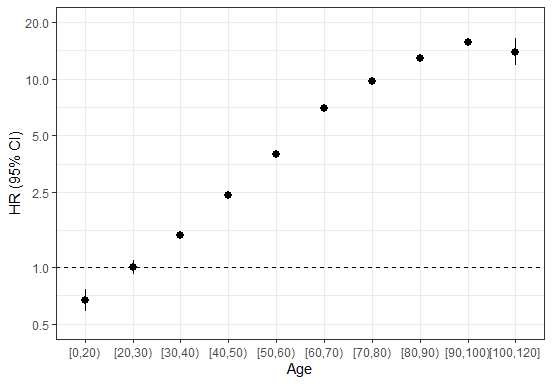

**Supplementary figure 15: Hazard ratios and 95% CIs for death per 10-year higher age by country** Model stratified by sex

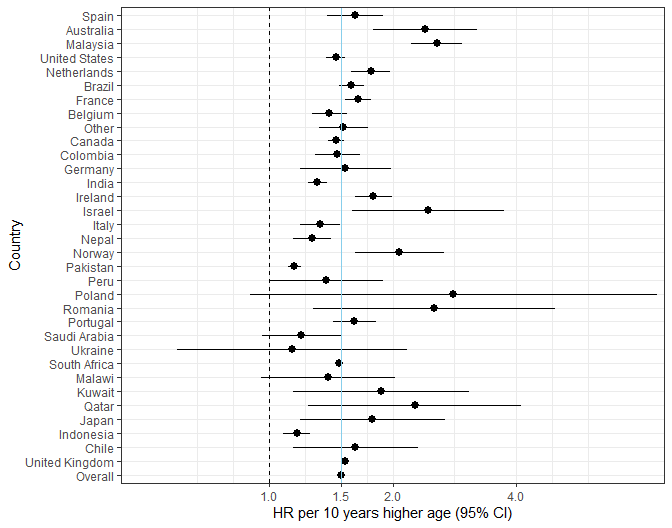

**Supplementary figure 16: Hazard ratios and 95% CIs for death for males vs females by country** Model adjusted for age (numeric)

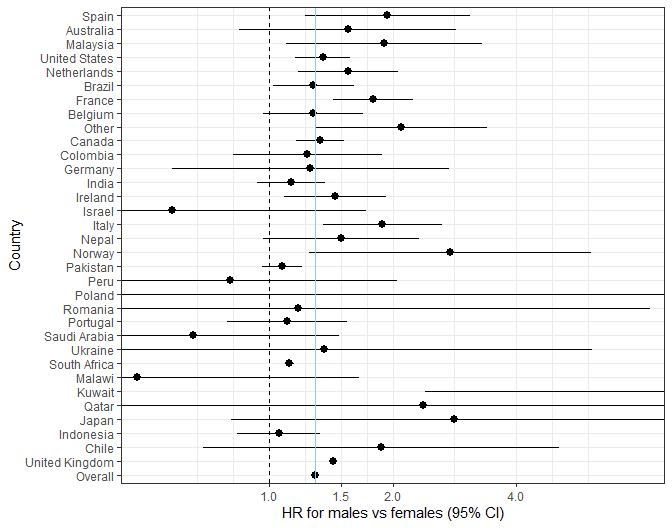

### Supplementary methods

**Case Report Forms**

The ISARIC-WHO Case Report Forms (CRFs) were used to collect data for individuals presenting with suspected or confirmed COVID-19. The CRFs are a part of the ISARIC/WHO Clinical Characterisation Protocol, a harmonised protocol for investigation of severe acute infections by pathogens of public health importance. Three versions of the CRF were available for use: the ISARIC WHO Clinical Characterisation Protocol for Severe Emerging Infections UK (CCP-UK) CRF, the COVID‐19 CORE CRF, and the COVID-19 RAPID CRF. The CCP-UK CRF was used to capture data for patients in UK hospitals, while the CORE and RAPID CRFs were used to capture data for patients outside the UK. The CORE CRF is a comprehensive form with variables collected to understand a broad spectrum of disease and management, while the RAPID CRF includes fewer variables than the CORE CRF and was designed for use where data collection resources are limited. CRFs were completed electronically through ISARIC’s Research Electronic Data Capture (REDCap®, version 8.11.11, Vanderbilt University, Nashville, Tenn.), hosted by the University of Oxford. Several regions did not use the ISARIC-WHO CRFs to collect data, but contributed data to the ISARIC data platform through their local CRFs or independent studies. All data submitted to the ISARIC data platform were harmonised to the CDISC SDTM standard (Study Data Tabulation Model) (version 1.7, Clinical Data Interchange Standards Consortium, Austin, Tex.).

For this study, data for 161,495 patients from 213 sites were collected using the CCP-UK CRF, 30,183 patients from 430 sites using the CORE CRF, and 5811 patients from 88 sites using the RAPID CRF. Other patient data were collected using locally-developed CRFs, usually based on the ISARIC CRFs.

**Variables used**

Age was categorized into 10-year groups, with age <20 and $\geq$ 100 into single groups.

Non-invasive ventilation was defined as BIPAP, CPAP or other unspecified type of non-invasive ventilation.

The following Covid-19 symptom definitions were used:

- World Health Organization (WHO):
  1. A combination of acute fever and cough,
  Or
  2. A combination of three or more of: fever, cough, general weakness and fatigue, headache, myalgia, sore throat, coryza, dyspnoea, anorexia, nausea and vomiting, diarrhoea, altered mental status
- Centers for Disease Control (CDC), United States:
  1. At least two of: fever, chills (not available), rigors (not available), myalgia, headache, sore throat, new olfactory and taste disorder,
  Or
  2. At least one of: cough, shortness of breath, difficulty breathing (not available)
- Public Health England
  New cough, or temperature $\geq$ 37.8 ${}^{\circ}$C, or a loss or change in sense of smell or taste
- European Centre for Disease Prevention and Control
  At least one of: cough, fever, shortness of breath, sudden onset anosmia, ageusia or dysgeusia
